# Life-course comorbidity patterns and integrated prediction of postpartum depression, multimorbidity, and symptom progression

**DOI:** 10.64898/2026.02.18.26346535

**Authors:** Selena Aranda, Ariadna Bada-Navarro, Bru Cormand, Marta Cano, Narcís Cardoner, Elisa Llurba, Marina Mitjans, Dora Koller

**Author notes:** Corresponding author: Dora Koller, PhD, Women and Perinatal Health Research Group, Institut de Recerca (IR) Sant Pau Carrer Sant Quintí, 77-79, 08025 Barcelona, Spain.

## Abstract

Perinatal depression (PD) is common and disabling, yet its longitudinal comorbidity patterns and predictability remain poorly understood. This study leveraged 8,804 women with delivery records in the All of Us cohort, including 438 with clinically diagnosed postpartum depression (PPD), to characterize multimorbidity trajectories and develop integrated prediction models. Comorbidities were grouped into 38 conditions across psychiatric, autoimmune, metabolic, neurological/pain, and reproductive/gynecological categories and examined both cross-sectionally and in monthly time bins from 250 months before to 500 months after delivery. Latent class analysis identified three pre- and post-delivery multimorbidity profiles and transitions between classes, while polygenic risk scores for depression and obstetric, clinical and socioeconomic variables were combined in machine learning models to predict PPD, post-delivery class membership, and symptom worsening among initially low-burden women. PPD cases showed higher odds of several psychiatric, autoimmune, and metabolic conditions and a tendency toward greater post-delivery comorbidity accumulation, particularly among women who were healthy pre-pregnancy. Multimorbidity profiles based on latent classes captured clinically meaningful risk gradients, and transition analyses revealed that incident PPD in previously healthy women marked a shift toward more symptomatic post-delivery profiles. Machine learning models achieved moderate discrimination for PPD and comorbidity outcomes and highlighted the importance of genetic liability, obstetric complications, and socioeconomic disadvantage, but low positive predictive values limit clinical implementation. These findings position PPD as a critical event in women’s psychiatric, cardiometabolic, and pain-related health trajectories and support life-course, multimorbidity-informed screening and prevention strategies that extend beyond the traditional postpartum period.

## INTRODUCTION

Perinatal depression (PD) is a form of major depressive disorder (MDD) occurring during the perinatal period, with a global prevalence of 17% (Wang et al., 2021), while postpartum depression (PPD) is its postpartum-onset form, representing 40.1% of PD cases (Wisner et al., 2013). The burden of PD is substantial, showing negative effects on mothers’ quality of life and long term physical and mental health (Dagher et al., 2021). From the economic perspective, the lifetime costs of PD were estimated at £75,728 per affected woman in the UK, and the aggregated costs were £6.6 billion (Bauer et al., 2016). The burden of PD extends beyond individual suffering, impacting families through strained relationships and compromised child development, with increased risks of behavioral, motor, language, and cognitive difficulties for the child (Fan et al., 2024). These effects highlight the need for increased resources to support women with PD and prevent long-term negative effects on both mothers and children.

PD is associated with substantial psychiatric and physical comorbidity burden that extends beyond the immediate postpartum period (Meltzer-Brody and Stuebe, 2014). Affected women face elevated risk for recurrent mood disorders, many developing chronic depression following an initial postpartum episode (Meltzer-Brody and Stuebe, 2014), and higher rates of anxiety, post-traumatic stress symptoms, and other mood disorders compared with women without PD (Slomian et al., 2019). PD significantly increases suicide risk, particularly within the first year after diagnosis with persistent risk elevations throughout the 18 years of follow-up (Yu et al., 2024), which is a leading cause of maternal mortality in high-income countries (Chin et al., 2022). Beyond psychiatric conditions, PD is consistently linked with a range of obstetric and metabolic complications, including higher rates of preterm birth, preeclampsia, and gestational diabetes, as well as greater likelihood of postpartum weight retention (Howard and Khalifeh, 2020). These associations suggest partially shared pathophysiological mechanisms involving dysregulated stress response, inflammation, and metabolic dysfunction, operating bidirectionally such that depression may precipitate pregnancy complications while obstetric events can reinforce depressive symptoms (Levin and Ein-Dor, 2023). Evidence further indicates that PD may serve as an early marker for long-term cardiometabolic risk, with metabolic syndrome and elevated lipid profiles persisting years beyond delivery (Lu et al., 2024). Despite these recognized associations, most existing studies are limited by modest sample sizes, cross-sectional or short-term designs, and a focus on isolated disorders rather than comprehensive multimorbidity patterns (Howard and Khalifeh, 2020; Watt et al., 2002; Zhang et al., 2025).

While some studies have identified symptom-based trajectory classes (M. Jiang et al., 2025) or examined heterogeneity in PD presentation (Postpartum Depression: Action Towards Causes and Treatment (PACT) Consortium, 2015), none have characterized broad comorbidity profiles spanning psychiatric, metabolic, autoimmune, and other physical conditions across both pre- and post-delivery periods, analyzed transitions between comorbidity classes, or examined symptom worsening trajectories.

Current prediction models for PD based on genetic data have very low prediction capacity (AUC = 0.52–0.56) (Byrne et al., 2014; Rantalainen et al., 2020; Tebeka et al., 2024). Machine learning models that rely solely on clinical and socioeconomic data have shown only modest predictive performance (Clapp et al., 2024; Reps et al., 2022), and studies reporting higher discrimination (AUC = 0.80, 0.88, and 0.91) relied on proxy outcomes such as screening scores, non-specific diagnostic codes, or antidepressant prescriptions rather than PD diagnosis (Munk-Olsen et al., 2022; Shin et al., 2020; Wakefield and Frasch, 2023). Machine learning methods offer the potential to integrate high-dimensional clinical, genetic, and socioeconomic information to capture complex, non-linear risk patterns, but have rarely been applied using validated PD outcomes in large, ancestrally diverse cohorts. Moreover, the lack of models that explicitly target symptom progression, comorbidity clustering, and transitions from low to high symptom burden limits the translational value of existing tools for early identification and prevention.

The present study leverages the large and diverse All of Us cohort to advance the understanding of the life-course trajectories of PPD. First, comorbidity patterns and temporal trajectories are characterized across pre-delivery and post-delivery periods, providing a longitudinal view of multimorbidity around childbirth. Second, latent class analysis is used to identify distinct comorbidity-based symptom profiles and to examine transitions between classes from pregnancy to the postpartum period. Third, integrated prediction models are developed that combine clinical, obstetric, socioeconomic, and polygenic risk information to predict PD, post-delivery comorbidity profiles, and symptom worsening among initially low-symptom women. Together, these aims are designed to improve personalized risk assessment, refine phenotypic stratification, and inform targeted preventive and early intervention strategies for women at risk of adverse perinatal mental health trajectories.

## METHODS

### Study participants

Data were obtained from the All of Us (AoU) Research Program (Controlled Tier, version 8), a large longitudinal cohort of over 633,000 adults in the United States designed to enhance the representation of historically underrepresented populations. We analyzed data from 8,804 female participants with electronic health record (EHR) data of a delivery event (“delivery finding”; SNOMED CT: 118215003). Among these participants, 438 were classified as PPD cases (SNOMED CT: 58703003), defined as PD with onset after delivery, while 8,366 participants had delivery information but no recorded diagnosis of PD and were therefore used as controls. In case of multiple deliveries, the first was included in the study. As all PD cases were identified during the postpartum period, they are hereafter referred to as PPD.

Additional data were collected for these participants, including age at recruitment and body mass index (BMI); obstetric characteristics (age at first delivery, number of pregnancies, cesarean delivery, twin pregnancy, preterm delivery, hypertension during pregnancy, gestational diabetes, and preeclampsia); and socioeconomic indicators (median household income, assisted income, high school education, lack of health insurance, poverty, vacant housing, and a composite deprivation index). Comorbid conditions were ascertained from EHR data and grouped into 38 diseases across five categories: reproductive/gynecological (n=5), psychiatric (n=15), autoimmune (n=8), neurological/pain/systemic (n=6), and metabolic/cardiometabolic (n=4) diseases (**Supplemental Table 1**). Prevalence of each variable was compared between PPD cases and controls using a t-test for continuous variables and a chi-square test for binary variables. Details about genotyping is described elsewhere (All of Us Research Program Genomics Investigators, 2024).

Analyses were conducted in the All of Us Researcher Workbench cloud environment using RStudio (version 4.4.0) and Python (via Jupyter Workbench) for statistical analyses.

### Comorbidity analyses

We compared the difference in odds of each of the 38 comorbidities at any time point between PPD cases and controls using logistic regression models. Odds ratios (ORs) and 95% confidence intervals (CIs) were estimated for each comorbidity, adjusting for age at delivery, BMI, and deprivation index. We also ran the analysis without BMI as a covariate, given that BMI is associated with higher prevalence of many diseases and has a bidirectional relationship with PPD, to compare differences between the two models. To account for multiple testing we applied false discovery rate (FDR) correction (q < 0.05).

To examine temporal patterns of comorbidity occurrence relative to delivery, we calculated the prevalence of each comorbidity in PPD cases and controls across monthly time bins spanning from 250 months before delivery to 500 months after delivery. For each comorbidity at each time point, we tested the difference in prevalence between PPD cases and controls using chi-square tests (or Fisher’s exact test when expected cell counts were <5). We applied FDR correction separately within each comorbidity across all time points examined to identify specific time windows with significantly different prevalence patterns. We also calculated mean prevalence differences for pre-delivery (months <0) and post-delivery (months ≥0) periods to characterize temporal patterns.

### Latent Class Analysis and post-analyses

Latent class analysis (LCA) was performed separately for the pre-delivery and post-delivery periods to identify clinically meaningful subgroups based on comorbidity patterns. Pre-delivery comorbidities were defined as those registered before date at first delivery, and post-delivery comorbidities were defined as those after the date at first delivery. Models with 2 to 5 latent classes were estimated independently for the pre-delivery and post-delivery datasets. Best model selection was based on the Akaike information criterion (AIC), Bayesian information criterion (BIC), entropy, clinical interpretability, and a minimum class size restriction (≥1% of the sample). Following model selection, individuals were assigned to their most likely latent class using maximum posterior probability (PP). Item-response probabilities were examined to characterize the comorbidity profile of each class. All latent class analyses were conducted using the poLCA package (version 1.6.0.1). To quantify the association between latent class membership and PPD, PPD prevalence was calculated within each pre-delivery and post-delivery class. Pairwise comparisons between classes were performed using Fisher’s exact test, with class 1 (minimal comorbidity) serving as the reference category.

To examine stability and change in comorbidity patterns across the perinatal period, a transition matrix was constructed among women with valid latent class assignments at both time points (N=8,680). The cross-tabulation of pre-delivery (rows) by post-delivery (columns) classes was summarized as counts and row-wise percentages, representing the probability of transitioning from each pre-delivery class to each post-delivery class. Diagonal cells represented stability (remaining in the same class profile), while off-diagonal cells represented transitions to different comorbidity patterns. The transition matrix was subsequently stratified by PPD status to evaluate whether comorbidity transitions differed between groups. Within each pre-delivery class, chi-square tests were used to compare the distribution of post-delivery classes between women with and without PPD.

### Polygenic risk scoring

In the full pregnancy cohort, 7,460 individuals had genetic data, of whom 357 had PPD and 7,103 were controls. The analytical sample was restricted to individuals genetically inferred as African (AFR), Admixed American (AMR), or European (EUR) ancestry, given adequate sample sizes in these groups (n_AFR_=1,939; n_AMR_=2,618; n_EUR_=2,697; n_total_=7,254, 339 PPD cases and 6,915 controls). Genotype data were quality-controlled using standard filters (Choi et al., 2020), retaining variants with minor allele frequency >1%, genotype call rate >95%, and Hardy-Weinberg equilibrium p>1×10-6.

The final target dataset included n_AFR_=1,665 (70 cases), n_AMR_=2,342 (89 cases), and n_EUR_=2,507 (155 cases) individuals after accounting for relatedness. Reference LD panels corresponding to AFR, AMR, and EUR ancestries from the 1000 Genomes Project were used for LD modeling (1000 Genomes Project Consortium et al., 2015). Summary statistics for major depression (MD) were used for each discovery ancestry (AFR: 36,818 MD cases and 161,679 controls (Meng et al., 2024), AMR: 25,013 MD cases and 352,946 controls (Meng et al., 2024), and EUR: 688,808 MD cases and 4,364,225 controls (Major Depressive Disorder Working Group of the Psychiatric Genomics Consortium, 2025)). We used PRS-CSx to transform effect sizes via inverse-variance weighted meta-analysis across ancestry-specific posterior effects, generating a single set of variant weights optimized for cross-ancestry prediction with the phi auto setting (Ruan et al., 2022). Depression PRS were then computed using PLINK2 (Chang et al., 2015), and the PRS estimates were standardized within each ancestry group. Age at delivery and the top-10 within-ancestry PCs were added as covariates.

### Machine learning analyses

The machine learning feature set included standardized PRS, genetic ancestry (AFR, AMR, and EUR), age at first delivery, BMI, number of pregnancies, pregnancy complications (cesarean section, twin pregnancy, preterm birth, hypertensive disorder, gestational diabetes, preeclampsia), socioeconomic indicators, and race. Categorical variables (race, ancestry) were label encoded and remaining predictors were kept numeric. Individuals without PRS were excluded from ML analyses, as genetic scores cannot be meaningfully imputed without genotype data. Median imputation was applied to continuous predictors with residual missingness (BMI and socioeconomic variables). All analyses included participants with complete PRS data and non-missing outcome and predictor variables (n=6,470).

We used a consistent 70/30 stratified train-test split for cross-validation. Due to class imbalance across all outcomes, models were fitted with class balancing: logistic regression with L2 regularization and balanced class weights, random forest with balanced class weights, and XGBoost with scale_pos_weight set to the ratio of majority to minority class. Five fold stratified cross validation on training data estimated balanced accuracy. Test set performance was evaluated using balanced accuracy, AUROC, sensitivity, specificity, positive predictive value (PPV), negative predictive value (NPV), and F1 score. Decision thresholds were optimized by varying cutoffs from 0.10 to 0.90 to maximize balanced accuracy on the test set. The model with the highest AUROC was selected as the best performer. Feature importance was assessed using the Gini importance metric from the best-performing tree-based model.

We performed three prediction analyses: 1) Prediction of PPD status (n=6,470; PPD cases n=308, 4.76%) using a training set of 4,529 individuals and test set of 1,941 with balanced case proportion; 2) Prediction of post-delivery comorbidity class: Using the same dataset (n=6,470), we predicted symptomatic profiles (latent classes 2 and 3 combined, 34.27%) versus minimal symptoms (latent class 1, 65.73%). To assess the predictive utility of PPD diagnosis, we trained two model sets, one without PPD as a predictor (baseline feature set), and with PPD included as a binary predictor; and 3) Prediction of symptom worsening: Among women with minimal symptoms (class 1) during pre-pregnancy (n=6,421), we predicted postpartum worsening (transition to classes 2 or 3, n=1,707, 26.6%) versus remaining stable in class 1 (n=4,714, 73.4%). We also evaluated models without and with PPD as a predictor.

## RESULTS

### Comorbidity associations

Characteristics of variables used in this study and prevalence differences in individuals with PPD and controls are shown in **Supplemental Table 2**. After adjusting for age at delivery, BMI, and socioeconomic deprivation, 11 of 38 comorbidities were associated with significantly higher odds among PD cases compared with controls (p < 0.05), of which six remained significant after FDR correction (q < 0.05, **Figure 1**, **Supplemental Table 3**). These included four psychiatric phenotypes, such as depressive disorder (OR=4.94, 95% CI: 3.89-6.27), anxiety disorder (OR=3.22, 95% CI: 2.58-4.02), Post-Traumatic Stress Disorder (PTSD; OR=1.64, 95% CI: 1.23-2.20), and Premenstrual Dysphoric Disorder (PMDD; OR=3.31, 95% CI: 1.38-7.92); as well as celiac disease (OR=4.11, 95% CI: 2.12-7.96) and Polycystic Ovary Syndrome (PCOS; OR=2.39, 95% CI: 1.67-3.44). In the model without BMI as a covariate, 9 comorbidities showed significantly higher odds among PPD cases compared with controls, all of which remained significant after FDR correction (**Supplemental Table 4**). The three additional phenotypes included migraine (OR=1.33, 95% CI: 1.08-1.64), irritable bowel syndrome (OR=1.56, 95% CI: 1.13-2.17), and chronic pain (OR=1.30, 95% CI: 1.06-1.59).

**Figure 1.**
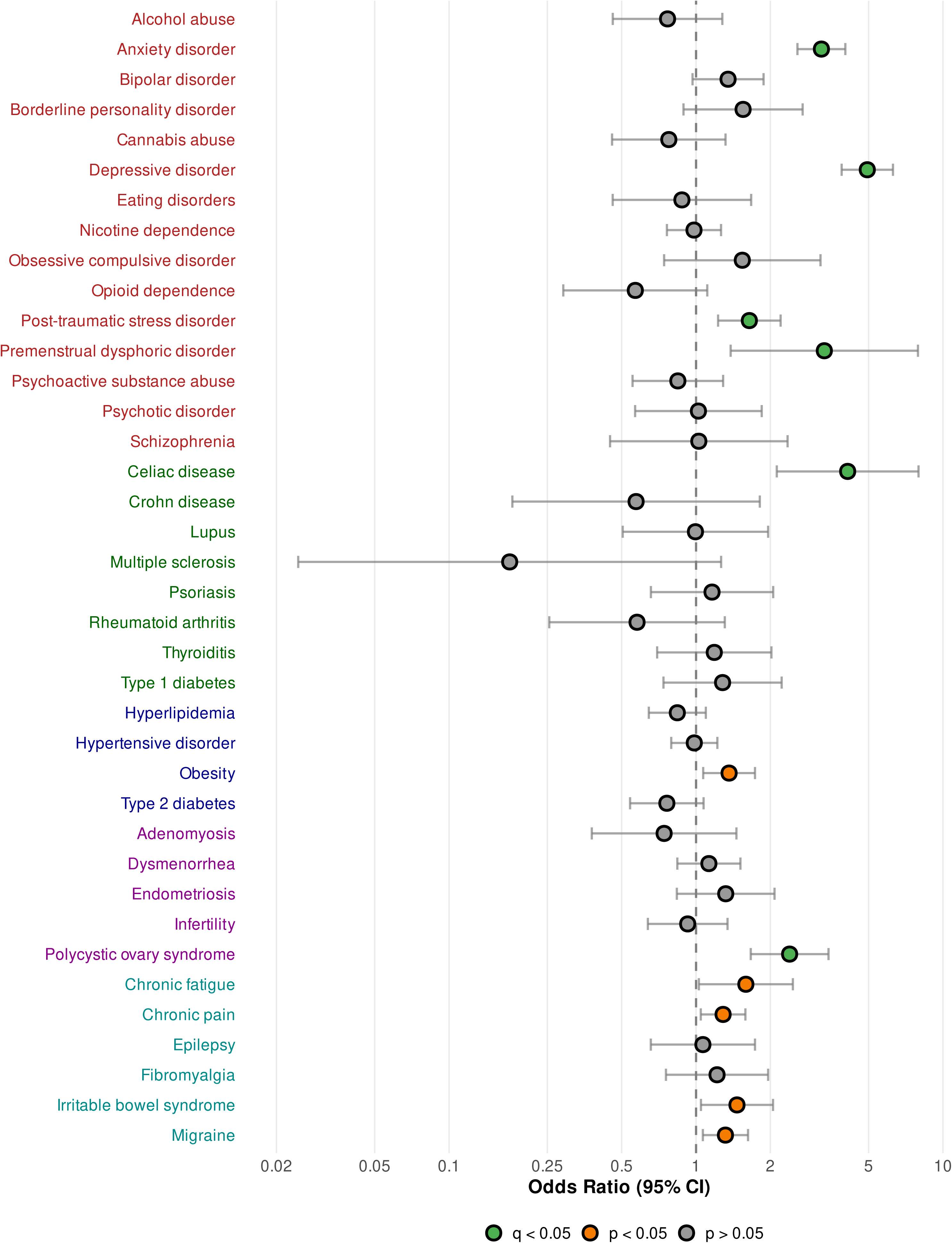
Association between postpartum depression and comorbid conditions. Forest plot showing odds ratios (ORs) and 95% confidence intervals (CIs) for the association between postpartum depression cases and controls across 38 comorbid conditions. All associations were estimated using logistic regression models adjusted for age at delivery, body mass index, and socioeconomic deprivation. Comorbidity names are colored by clinical category: psychiatric (red), autoimmune (dark green), metabolic/cardiometabolic (dark blue), reproductive/gynecological (purple), and neurological/pain/systemic (teal). Points are colored by statistical significance: green indicates false discovery rate (FDR)-corrected significance (q < 0.05), orange indicates nominal significance (p < 0.05), and grey indicates non-significant associations (p > 0.05). The dashed vertical line indicates an OR of 1.0 (no difference between cases and controls). ORs greater than 1 indicate higher odds of the comorbidity in postpartum depression cases compared to controls.

### Temporal trajectory of comorbidities

Examination of temporal patterns revealed that 7 of the 38 comorbidities exhibited significantly different prevalence between PPD cases (N=438) and controls (N=8,366) at specific time points after FDR correction (**Figure 2**, **Supplemental Figure 1**, **Supplemental Table 5**). Psychiatric conditions dominated these temporal patterns, with depressive disorder showing significant differences at 5 time points (months 0, 1, 2, 3 and 29 post-delivery; peak difference of 4.1% at delivery, p=1.91×10^-7^) and anxiety disorder at 4 time points (months -17, 0, 2, and 4; peak difference of 2.8% at delivery, p=6.64×10^-6^). PTSD exhibited a single significant time point at month 19 post-delivery (difference 0.7%, p=1.22×10^-4^). Among neurological/pain/systemic conditions, chronic pain showed significant differences at 2 time points (month 27 pre-delivery (difference=0.89%, p=8.37×10^-5^) and month 5 post-delivery (difference=0.96%, p=3.73×10^-5^)), while chronic fatigue (month 5, difference=0.69%, p=1.22×10^-4^) and fibromyalgia (month 77, difference=0.69%, p=1.22×10^-4^) each showed single significant time points. Hypertensive disorder exhibited a single significant pre-delivery time point at month 4 (difference=1.33%, p=1.99×10^-6^) (**Figure 2**, **Supplemental Figure 1, Supplemental Table 5**).

**Figure 2.**
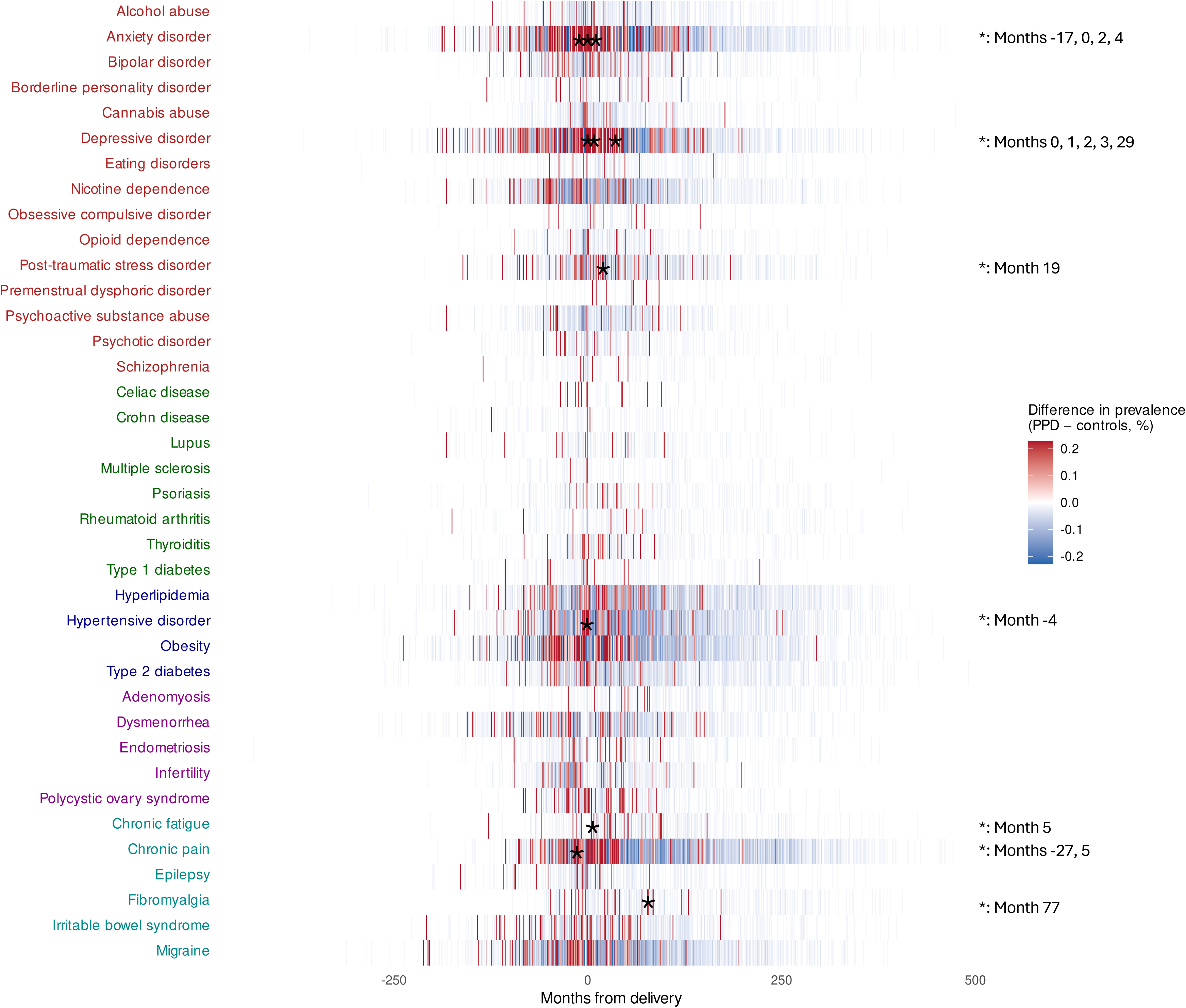
Temporal trajectories of comorbidity prevalence differences in postpartum depression Heatmap showing the difference in comorbidity prevalence between postpartum depression (PPD) cases and controls across time relative to delivery. Each tile represents the absolute percentage-point difference (PPD cases minus controls) for a given comorbidity at each monthly time bin. Red shades indicate higher prevalence in PPD cases, blue shades indicate higher prevalence in controls, and white indicates no difference. Comorbidities are grouped by clinical category (indicated by text color) and ordered alphabetically within each category: psychiatric (red), autoimmune (dark green), metabolic/cardiometabolic (dark blue), reproductive/gynecological (purple), and neurological/pain/systemic (teal).

Temporal patterns revealed a consistent trend of higher mean prevalence differences in the post-delivery period compared to the pre-delivery period across 30 out of the 38 comorbidities. These spanned all disease categories, with top results of chronic pain in the neurological/pain/systemic group (prevalence_pre-delivery_=5.9%, prevalence_post-delivery_=27.7, p=4.4×10^-273^), hyperlipidemia in the metabolic/cardiometabolic category (prevalence_pre-delivery_=5.7%, prevalence_post-delivery_=16, p=4.4×10^-273^), anxiety disorder in the psychiatric category (prevalence_pre-delivery_=16.6%, prevalence_post-delivery_=29.7, p=1.53×10^-73^), adenomyosis in the reproductive/gynecological category (prevalence_pre-delivery_=0.4%, prevalence_post-delivery_=2.6, p=5.14×10^-33^), and rheumatoid arthritis in the autoimmune category (prevalence_pre-delivery_=0.6%, prevalence_post-delivery_=2, p=1.21×10^-14^) (**Supplemental Table 6**).

### Latent Class Analysis

Overall, three-class solutions were retained for both the pre,- and post-delivery periods as they captured substantial improvements in model fit while maintaining good class separation and interpretability, whereas additional classes yielded minimal fit gains and reduced classification quality (**Supplemental Table 7**).

For the pre-delivery dataset, the three classes were defined as minimal/low comorbidity burden (class 1) with 74.9% of the cohort, moderate multimorbidity (class 2) with 19.8% of the cohort (most common comorbidities: depressive disorder, anxiety disorder, hypertensive disorder, migraine and obesity), and high multimorbidity (class 3) with 5.3% of the cohort (most common comorbidities: depressive disorder, anxiety disorder, nicotine dependence, psychoactive substance abuse, bipolar disorder, and obesity) (**Supplemental Table 8**). When comparing the class distribution by PPD status, we found that significantly less PPD cases belonged to the minimal comorbidity group compared to controls (55.9% vs 75.8%, respectively, p=7.85×10^-21^), more PPD cases belonged to the moderate multimorbidity group (35.2% vs 19%, respectively, p=2.66×10^-16^) and to the high comorbidity group (8.9% vs 5.1%, respectively, p=8.56×10^-4^) (**Figure 3**). Furthermore, when comparing PPD prevalence between classes, PPD prevalence was higher in the moderate multimorbidity class compared to the minimal class (8.9% vs 3.8%, respectively, p=1.21×10^-18^), and in the high multimorbidity class compared to the minimal class (8.4% vs 3.8%, respectively, p=1.61×10^-6^) (**Supplemental Table 9**).

**Figure 3.**
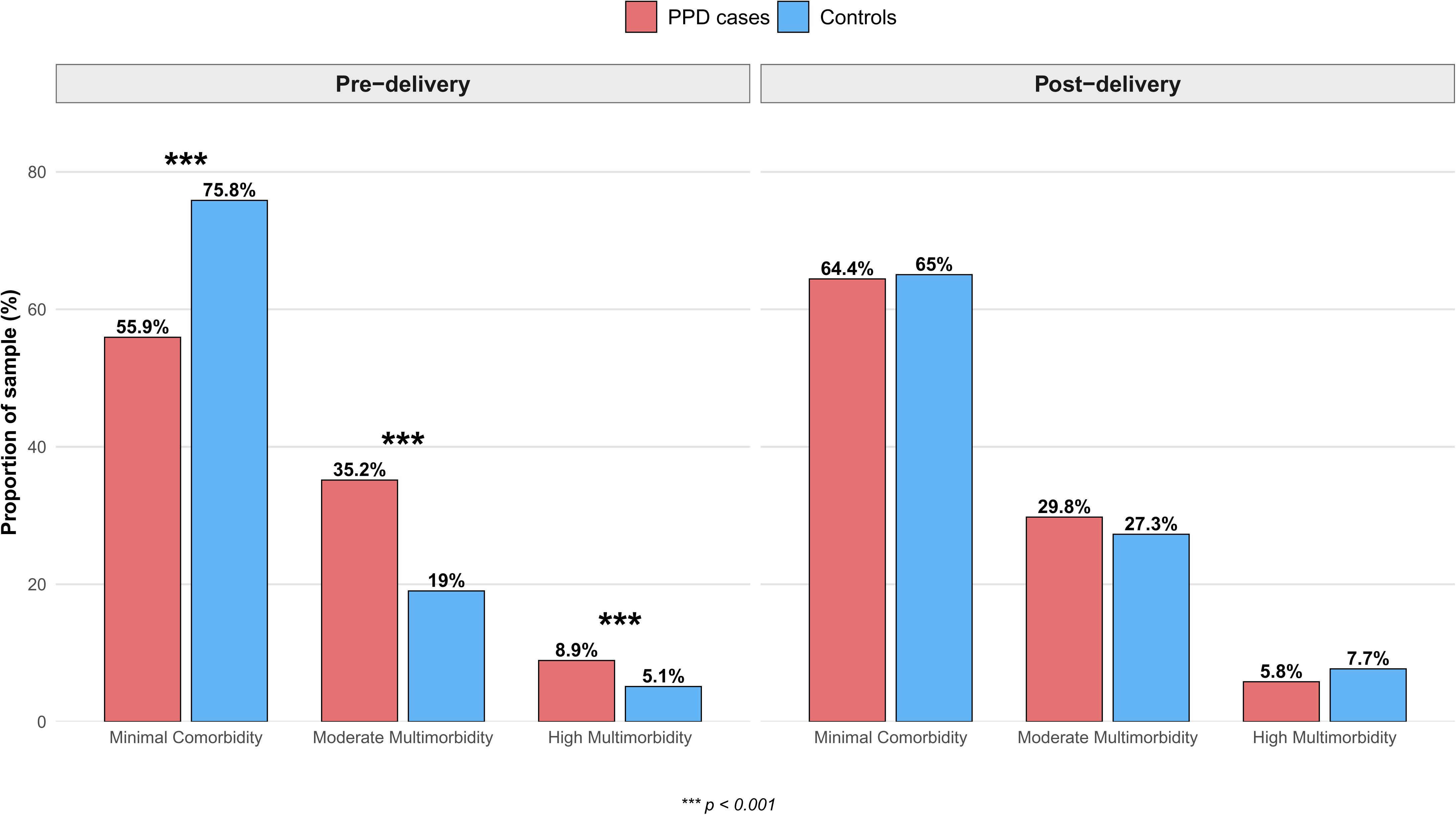
Latent class proportions in PPD cases and controls. Distribution of comorbidity-based latent classes in women with postpartum depression (PPD) versus controls during pre-delivery (left) and post-delivery (right) periods. Pre-delivery, PPD cases showed significantly higher multimorbidity burden with fewer women in the minimal comorbidity class and more in moderate and high multimorbidity classes (all p<0.001). Post-delivery, class distributions were similar between PPD cases and controls.

For the post-delivery dataset, the three classes were defined similarly to the pre-delivery dataset, as minimal/low comorbidity burden (class 1) with 65% of the cohort, high multimorbidity (class 2) with 7.6% of the cohort (most common comorbidities: depressive disorder, anxiety disorder, nicotine dependence, post-traumatic stress disorder, bipolar disorder, migraine, chronic pain, hypertensive disorder, and obesity), and moderate multimorbidity (class 3) with 27.4% of the cohort (most common comorbidities: depressive disorder, anxiety disorder, chronic pain, hypertensive disorder, migraine, and obesity) (**Supplemental Table 10**). When comparing the class distribution by PPD status, we found no difference between PPD cases and controls (minimal group: 64.4% vs 65%, respectively, p=0.83; moderate multimorbidity group: 29.8% vs 27.3%, respectively, p=0.28); and high multimorbidity group: 5.8% vs 7.7%, respectively, p=0.18 (**Figure 3**). Furthermore, when comparing PPD prevalence between classes, no difference was found either (**Supplemental Table 9**).

### Transitions in comorbidity class membership across the perinatal period

Among women with valid latent class assignments at both pre- and post-delivery time points (N=8,680), substantial stability in comorbidity patterns was observed, with most women transitioning to or remaining in the minimal/low comorbidity class post-delivery (**Figure 4**, **Supplemental Table 11**).

**Figure 4.**
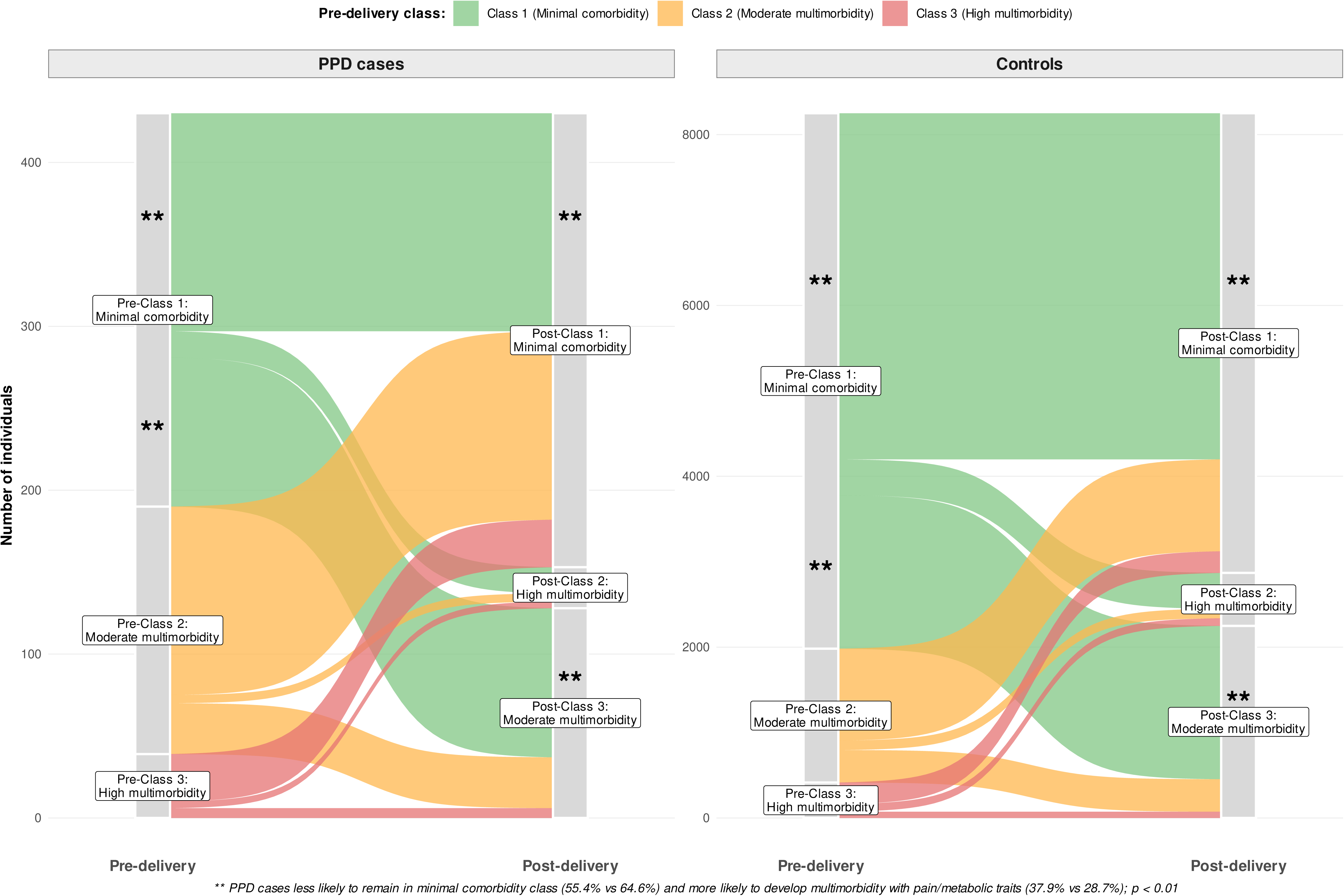
Class transitions in PPD cases and controls. Transitions between comorbidity-based latent classes from pre-delivery to post-delivery periods in postpartum depression (PPD) cases (left) and controls (right). Among women with minimal comorbidity pre-delivery, PPD cases were significantly less likely to remain in the minimal class post-delivery and more likely to transition to moderate multimorbidity compared with controls (both p<0.01). Transition patterns were similar between PPD cases and controls for women starting in moderate or high multimorbidity classes.

Among women classified in the minimal/low comorbidity class pre-delivery (pre-class 1), 64.3% remained in the same class post-delivery, 29% transitioned to the moderate multimorbidity class, and 6.7% transitioned to the high multimorbidity class. Among women with moderate multimorbidity pre-delivery (pre-class 2), 69.3% transitioned to the minimal comorbidity class post-delivery, 23.9% remained in the moderate multimorbidity class, and 6.8% transitioned to the high multimorbidity class. Among women with high multimorbidity pre-delivery (pre-class 3), 62.1% transitioned to the minimal comorbidity class post-delivery, 17.4% transitioned to the moderate multimorbidity class, and 20.5% remained in the high multimorbidity class (**Supplemental Table 11**). Among women in the minimal/low comorbidity class pre-delivery (pre-class 1), differences in post-delivery class membership by PPD status were observed.

Women with PPD were less likely than controls to remain in the minimal comorbidity class post-delivery (55.4% vs. 64.6%, p=4.23×10^-3^), and a greater proportion of women with PPD transitioned to the moderate multimorbidity class post-delivery compared with controls (37.9% vs. 28.7%, p=2.43×10^-3^), meaning that PPD cases were at higher risk of developing new comorbidities and less likely to maintain their relatively healthy status. The proportion transitioning to the high multimorbidity class post-delivery was identical between women with and without PPD (6.7% in both groups, *p* = 1). Furthermore, no differences were found when transitioning from the moderate and high multimorbidity classes to any other class (**Figure 4**, **Supplemental Table 11**).

### Machine learning prediction models

#### PPD prediction

Random forest with class balancing showed the best overall discrimination, with AUROC 0.72 on the test set (**Figure 5**). At the default 0.5 threshold, the model showed high specificity (0.99) and very low sensitivity (0.12), with a balanced accuracy of 0.55. Optimizing the threshold to 0.24 improved sensitivity to 0.57 and specificity to 0.77, with a balanced accuracy of 0.67, PPV 0.11, NPV 0.97, and F1-score 0.18 (**Supplemental Figure 3**, **Supplemental Table 12**). At the optimized threshold, the model correctly identified 52 of 92 women (56.5%) with PPD while accurately classifying 1414 (76.5%) women who did not develop PPD. The low PPV (33.9%) indicates that approximately 1 in 3 women flagged as high-risk actually developed PPD, while the high NPV (91.3%) demonstrates that more than 9 in 10 controls were correctly classified (**Supplemental Table 13**). The most important features in the prediction model were age at first delivery (0.15), PRS (0.13), and BMI (0.11) (**Supplemental Figure 3E**). Metrics from XGBoost and logistic regression are shown in **Supplemental Figure 3** and **Supplemental Table 2**.

**Figure 5.**
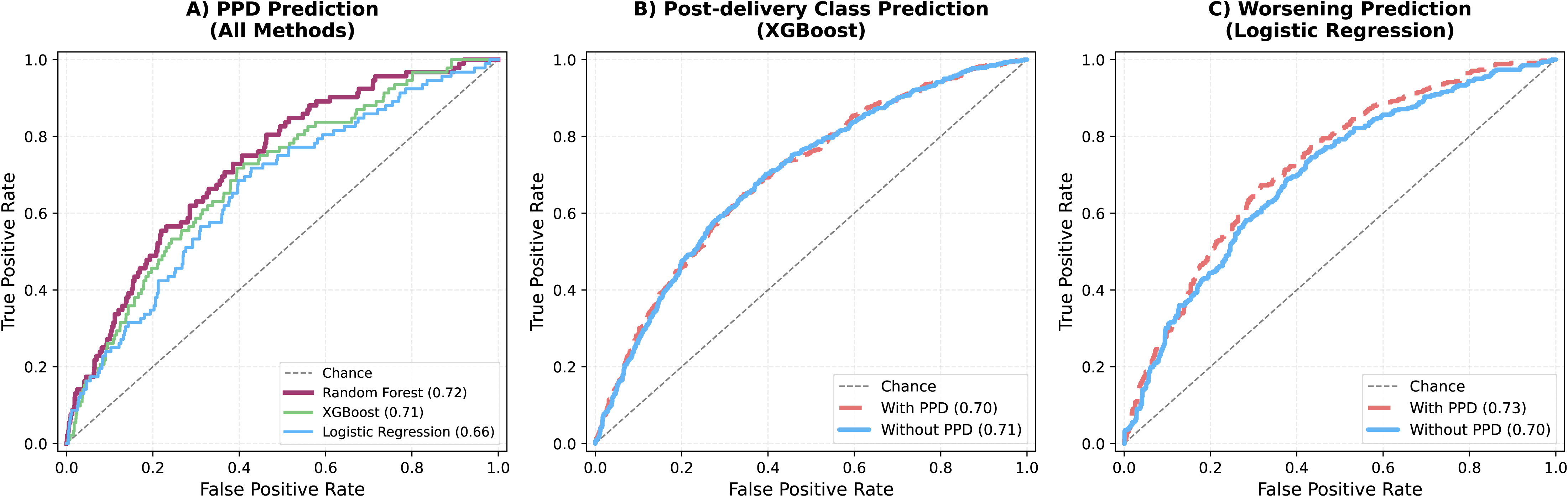
Machine learning prediction results. Predictors were pregnancy complications, socioeconomic factors, age at delivery, polygenic risk score for major depression, and pre-delivery latent class. A) Prediction of postpartum depression. B) Prediction of post-delivery asymptomatic (class1) or symptomatic (class 2: high multimorbidity or class 3: moderate multimorbidity) classes with and without PPD as an additional predictor. C) Prediction of symptom worsening (transitioning from pre-delivery class 1 (minimal comorbidity) to post-delivery symptomatic classes (class 2: high multimorbidity or class 3: moderate multimorbidity) with and without PPD as an additional predictor.

### Post-delivery comorbidity class prediction

XGBoost demonstrated the highest AUROC (0.72) (**Figure 5**) and, after threshold optimization to 0.53, achieved balanced accuracy 0.66 with sensitivity 0.53, specificity 0.79, PPV 0.57, and NPV 0.76 (**Supplemental Figure 4**, **Supplemental Table 14**). At the optimized threshold, XGBoost correctly identified 355 of 665 symptomatic women (53.4%) while maintaining high specificity (78.6%), meaning it accurately classified nearly 8 in 10 women with minimal symptoms. The moderate PPV (56.5%) indicates that approximately half of women predicted to have symptomatic comorbidity profiles actually presented with such symptoms postpartum, while the NPV (76.4%) demonstrates that approximately 3 in 4 women predicted to have minimal symptoms were correctly classified (**Supplemental Table 15**). The most important features in the prediction model were no health insurance (0.09), preterm birth (0.08), pre-delivery latent class (0.06), and number of pregnancies (0.06) (**Supplemental Figure 4E**). Adding PPD as a predictor resulted in no changes in AUROC (ΔAUROC: -0.001 (logistic regression and random forest) to -0.019 (XGBoost)) (**Figure 5**, **Supplemental Table 14**).

### Class transitions from minimal to symptomatic classes (symptom worsening)

Logistic regression achieved the highest AUROC of 0.70 (**Figure 5**). After threshold optimization to 0.47, balanced accuracy improved to 0.66 with sensitivity of 0.74, specificity of 0.57, PPV of 0.39, and NPV of 0.86 (**Supplemental Figure 5**, **Supplemental Table 16**). The optimized threshold increased sensitivity from 0.68 to 0.74, correctly identifying 254 of 342 women (74.3%) who worsened postpartum. This improvement was achieved while maintaining a high negative predictive value (85.9%; 538 of 626 women classified as low risk did not experience symptom worsening), indicating strong performance in ruling out worsening.

However, the PPV remained low (39%), meaning that fewer than 4 in 10 women flagged as high risk actually worsened postpartum, resulting in a high false-positive rate (n=405) (**Supplemental Table 17**). The other two models showed comparable metrics, random forest showing AUROC 0.70 and balanced accuracy of 0.66 at optimized threshold 0.47, while XGBoost demonstrated AUROC of 0.68 and balanced accuracy of 0.65 at optimized threshold 0.43 (**Supplemental Table 16**). The most important coefficients in the prediction model were deprivation index (0.69), high school education (0.59), BMI (0.31), and number of pregnancies (0.28) (**Supplemental Figure 5E**). Adding PPD status substantially improved the prediction of worsening among initially asymptomatic women. Logistic regression with PPD achieved AUROC 0.73 (vs 0.70 without PPD; ΔAUROC +0.027, ΔBalanced accuracy +0.021), while XGBoost showed AUROC 0.72 (vs 0.68 without PPD; ΔAUROC +0.046, ΔBalanced accuracy +0.026) (**Figure 5**, **Supplemental Table 16**).

## DISCUSSION

With comprehensive comorbidity profiling and machine learning in a large, multi-ancestry cohort, this study identifies previously healthy women who develop PPD as a high-risk group for subsequent multimorbidity emergence, suggesting a critical window for preventive intervention. Beyond psychiatric comorbidities, significant associations with autoimmune, metabolic, and pain conditions point to shared pathophysiological mechanisms warranting integrated clinical approaches and challenging the conceptualization of PPD as an isolated psychiatric event.

PPD cases showed elevated odds of psychiatric conditions (depressive disorder, anxiety disorder, PTSD, PMDD), autoimmune disorders (celiac disease), and metabolic conditions (PCOS), consistent with established evidence of chronic comorbidity following PPD (Agius et al., 2016; Pereira et al., 2022; Schoretsanitis et al., 2022; Tortora, 2015), though the particularly strong association for celiac disease (OR=4.11) warrants replication. The emergence of significantly higher odds for migraine, IBS, and chronic pain only when BMI was excluded suggests these conditions share inflammatory and metabolic pathways with obesity in women with PPD (Payne and Maguire, 2019).

Temporal trajectory analysis revealed that women with PPD showed significantly higher prevalence of psychiatric comorbidities clustering around delivery compared to controls, with depressive and anxiety disorders exhibiting peak differences at delivery. These diagnostic codes refer to non-perinatal-specific ICD codes, highlighting that PPD is not reliably captured in the clinical diagnosis coding of EHRs, a limitation previously documented (Slezak et al., 2023).

Women with PPD showed elevated anxiety 17 months pre-delivery, confirming pre-existing vulnerability (Guintivano et al., 2018), while the persistence of depressive disorder differences at 29 months post-delivery indicates chronic depression trajectories following PPD onset, consistent with evidence that untreated PPD frequently becomes persistent or recurrent (Slomian et al., 2019). Chronic pain was significantly more prevalent among PPD cases at both over 2 years before pre-delivery (month -27) and recent post-delivery (month 5) timepoints. While pregnancy-related musculoskeletal pain (e.g., back pain (Long et al., 2020), pelvic pain (Halliday et al., 2024)) has been documented as both risk factor and consequence of PPD, our findings extend this to general chronic pain conditions independent of obstetric complications, suggesting broader pain-mood bidirectionality beyond pregnancy-specific mechanisms, similar to non-perinatal depressive disorders (Miller-Matero, 2024) . Chronic fatigue emerged as significantly elevated 5 months postpartum among women with PPD, potentially reflecting the compounding effects of sleep disruption (Leistikow and Smith, 2024), caregiving demands (Matsuda et al., 2021), anxiety comorbidity (Agius et al., 2016), and the neurovegetative symptoms characteristic of depression itself (Dean and Keshavan, 2017). The elevated prevalence of hypertensive disorder 4 months pre-delivery among women who subsequently developed PPD likely captures gestational hypertension and preeclampsia cases, which again reflects potential diagnostic miscoding (Chen et al., 2020). PTSD (month 19) and fibromyalgia (month 77, over 6 years postpartum) emerged significantly later among PPD cases. Across the entire cohort, 30 of 38 conditions showed higher prevalence post-delivery compared to pre-delivery, spanning all five disease categories. These patterns demonstrate that PPD occurs within a context of progressive multimorbidity accumulation across multiple physiological systems, extending years beyond delivery. This finding aligns with a recent UK Biobank study showing PPD was associated with 1.13-fold higher risk of multimorbidity and faster accumulative rates of chronic diseases in women’s mid-late life (Zhang et al., 2025), warranting comprehensive longitudinal monitoring rather than time-limited postpartum care.

The substantial overrepresentation of subsequent PPD cases in moderate and high multimorbidity classes pre-delivery in the LCA analysis suggests that comorbidity burden may be a more powerful predictor than individual risk factors assessed in isolation, aligning with growing recognition that multimorbidity assessment improves clinical risk prediction (Y. Jiang et al., 2025). The moderate class profile, characterized by mood, anxiety, vascular, and pain conditions, may reflect a stress-sensitive phenotype (Postpartum Depression: Action Towards Causes and Treatment (PACT) Consortium, 2015) with dysregulated hypothalamic-pituitary-adrenal axis and inflammatory pathways, while the high class profile featuring depression, anxiety, PTSD, substance use and bipolar disorder alongside obesity suggests more severe psychiatric vulnerability. The convergence of post-delivery class distributions between PPD cases and controls, alongside with the substantial proportion of women transitioning from moderate and high multimorbidity classes pre-delivery to minimal comorbidity post-delivery (69.3% and 62.1%, respectively) reflects three methodological considerations. First, many pre-delivery comorbidities are pregnancy-specific conditions that usually resolve after delivery, resulting in apparent improvement in comorbidity burden. Second, comorbidity classes were defined using period-specific diagnoses: pre-delivery classes captured conditions diagnosed before first delivery, while post-delivery classes captured conditions diagnosed after first delivery. This temporal separation means pre-delivery diagnoses do not automatically carry forward to post-delivery classification unless registered again. Third, EHR coding practices may contribute to apparent class transitions, as chronic conditions that are managed but not actively re-documented post-delivery (e.g., depression in remission, controlled hypertension) would not appear in post-delivery class assignment despite ongoing clinical relevance. However, these overall transition patterns mask critical individual heterogeneity, as class transition analyses revealed that initially healthy women with PPD showed disproportionate worsening trajectories. This worsening may be driven by mechanisms including chronic stress (Xia et al., 2016), systemic inflammation (Osborne and Monk, 2013), lifestyle disruption (Brunson et al., 2024), and reduced healthcare engagement following PPD onset (Whyler et al., 2024). In contrast, women with established pre-delivery multimorbidity showed similar transition patterns regardless of PPD status, suggesting more deterministic trajectories less influenced by acute postpartum mood episodes. These findings have important care implications: initially healthy women may benefit most from PPD prevention and early intervention to avert multimorbidity cascade, while women with pre-existing multimorbidity require integrated chronic disease management targeting clustered psychiatric, pain, and cardiometabolic conditions.

The PPD prediction model achieved only moderate discrimination and low positive predictive value, indicating that current ML approaches are better suited for risk stratification and ruling out low risk women than for confidently identifying who will develop PPD. This is consistent with prior work showing that, for relatively low-prevalence outcomes, even reasonably accurate models generate many false positives (Byrne et al., 2014; Clapp et al., 2024; Rantalainen et al., 2020; Tebeka et al., 2024). Age at first delivery, BMI, and depression PRS were the top predictors, demonstrating that genetic liability contributes meaningfully to PPD risk alongside clinical and sociodemographic factors. However, the moderate overall model performance (AUROC=0.72) likely reflects both the limited sample size (308 PPD cases in the genotyped cohort) and the reduced power of cross-ancestry PRS in non-European populations, suggesting that larger, more ancestrally diverse training samples will be necessary to fully realize the potential of integrated genetic-clinical prediction models. In predicting post delivery comorbidity profiles, the leading role of socioeconomic and obstetric factors (e.g. lack of health insurance, preterm birth, pre delivery class, number of pregnancies) reinforces that multimorbidity trajectories are strongly shaped by socioeconomic disadvantage and pregnancy complications, not only by psychiatric diagnosis (Moreno-Juste et al., 2023). The absence of added predictive value from including PPD status suggests that comorbidity risk is largely encoded in pre existing health and social context, suggesting the need of comorbidity focused screening beyond women with diagnosed PPD (Easter et al., 2019). For symptom worsening among initially low burden women, the improvement in prediction when PPD is included indicates that a PPD episode may function as a pivotal life-course event, signaling elevated risk for subsequent multimorbidity. The fact that deprivation, education, BMI, and parity also rank highly points to modifiable social and metabolic pathways through which PPD may translate into long term physical health consequences. In summary, these models illustrate that ML can support more nuanced risk stratification across perinatal mental and physical health but, at current performance levels, should be viewed as decision support rather than decision making tools.

This study leveraged the large, diverse All of Us cohort with comprehensive EHR-based phenotyping across 38 comorbidities spanning five clinical categories and longitudinal assessment from 250 months pre-delivery to 500 months post-delivery. The integration of clinical, socioeconomic, and genetic data through machine learning, novel latent class transition analysis, and multi-ancestry PRS approach represent methodological advances addressing gaps in perinatal research. However, EHR-based phenotyping may under-ascertain conditions due to healthcare access barriers, non-perinatal-specific diagnostic coding, and stigma, potentially underestimating true comorbidity burden. The sample was restricted to women with documented deliveries and EHR data, potentially biasing toward populations with greater healthcare engagement and more complex medical needs. Cross-sectional comorbidity assignment precludes causal inference, and the observational design cannot determine directionality of associations. Limited sample size (438 PPD cases, of which 308 genotyped) restricted power for ancestry-stratified analyses and contributed to modest prediction performance. Machine learning models showed moderate discrimination (AUROCs 0.70-0.73) with low positive predictive values, limiting immediate clinical translation. Use of major depression PRS as a proxy for PPD-specific genetic risk may not fully capture perinatal-specific architecture.

## Data Availability

All data produced in the present work are contained in the manuscript.

## ACKNOWLEDGMENTS

The authors acknowledge support from ’Fundació La Marató de TV3’ (202218-31 to B.C.), the Spanish ‘Ministerio de Ciencia, Innovación y Universidades’: projects PID2021-1277760B-I100 and PID2024-158634OB-I00 funded by MICIU/AEI/10.13039/501100011033/ and FEDER-EU to B.C.; PID2022-139740OA-I00 funded by MICIU/AEI/10.13039/501100011033/ and FEDER-EU; RYC2021-033573-I funded by MCIN/AEI/10.13039/501100011033 and by the European Union “NextGenerationEU”/PRTR to M.M.; RYC2024-050099-I and JDC2024-055161-I funded by MICIU/AEI/10.13039/501100011033 and by FSE+ to D.K. and S.A., respectively, and AGAUR (2021SGR-01093 to B.C. and M.M.).

We also acknowledge the contribution of the participants and the investigators involved in the AoU Research Program. The program is supported by the National Institutes of Health, Office of the Director: Regional Medical Centers: 1 OT2 OD026549; 1 OT2 OD026554; 1 OT2 OD026557; 1 OT2 OD026556; 1 OT2 OD026550; 1 OT2 OD 026552; 1 OT2 OD026553; 1 OT2 OD026548; 1 OT2 OD026551; 1 OT2 OD026555; IAA: AOD 16037; Federally Qualified Health Centers: HHSN 263201600085U; Data and Research Center: 5 U2C OD023196; Biobank: 1 U24 OD023121; The Participant Center: U24 OD023176; Participant Technology Systems Center: 1 U24 OD023163; Communications and Engagement: 3 OT2 OD023205; 3 OT2 OD023206; and Community Partners: 1 OT2 OD025277; 3 OT2 OD025315; 1 OT2 OD025337; 1 OT2 OD025276.

## AUTHOR CONTRIBUTIONS

Conceptualization: D. K.; methodology: S.A., A.B., M.M., D.K.; analysis: S.A., D.K.; writing-original draft preparation: D.K.; writing-review and editing: all authors; supervision: D.K. All authors have read and agreed to the published version of the manuscript.

