## Supplemental Figures for "Life-course comorbidity patterns and integrated prediction of postpartum depression, multimorbidity, and symptom progression"

**Supplemental Figure 1**. Temporal trajectories of comorbid conditions in women with postpartum depression and controls.

Each panel shows one comorbidity with evidence of a significant temporal association with perinatal depression, including anxiety disorder, depressive disorder, hypertensive disorder, chronic pain, chronic fatigue, fibromyalgia, and post-traumatic stress disorder. Prevalence (percentage of participants with the comorbidity) is plotted across months relative to delivery, with separate lines for perinatal depression cases (red) and controls (blue).

The vertical dashed line at 0 marks the month of delivery. Grey shaded regions denote months in which the between-group difference in prevalence is statistically significant after false discovery rate correction ($q < 0.05$). Yellow points on the red line highlight specific time points for perinatal depression cases at which the comorbidity prevalence differs significantly from controls ($q < 0.05$).

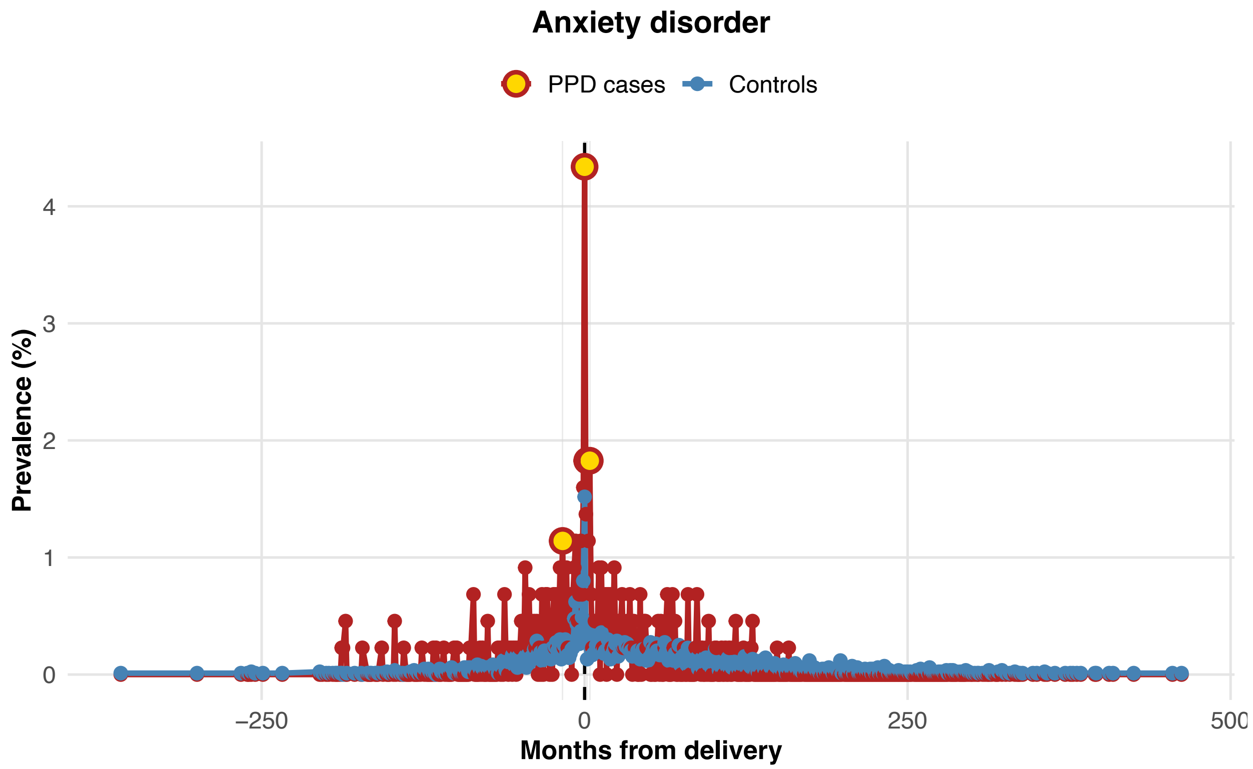

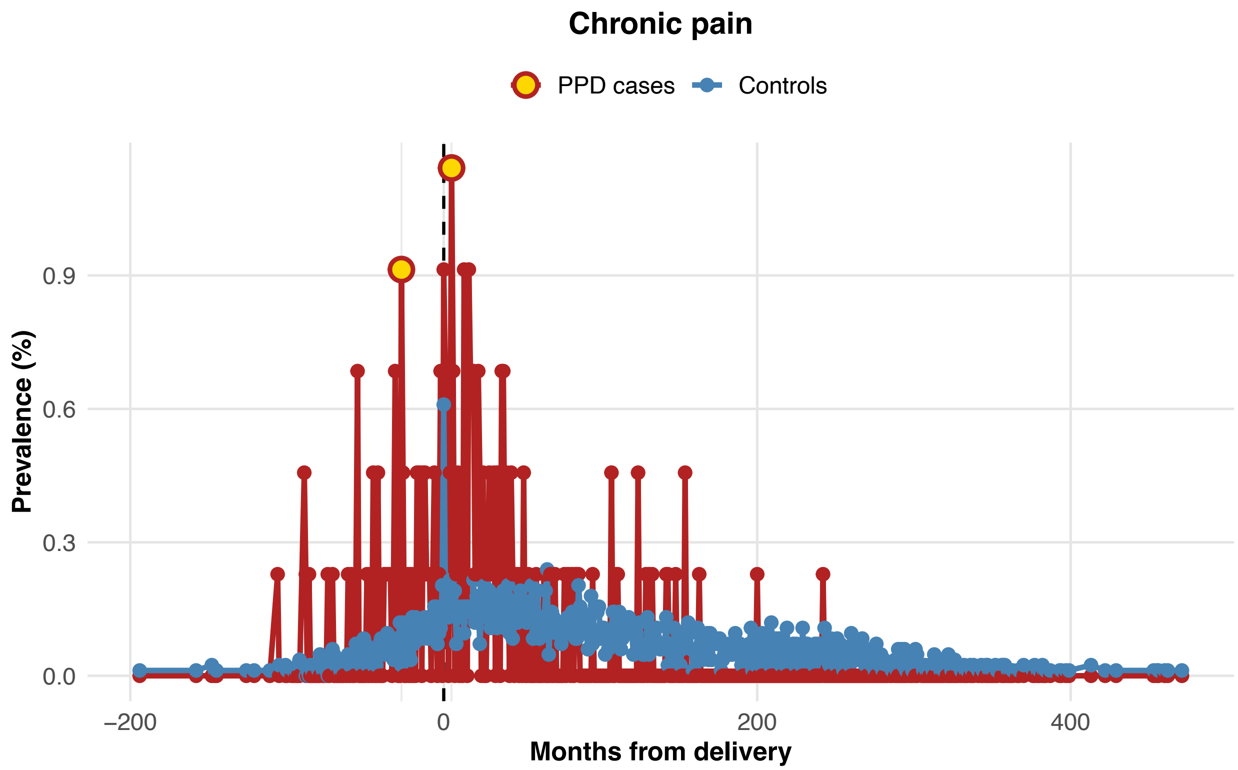
Chronic pain
PPD cases
Controls
Prevalence (%)
0.0
0.3
0.6
0.9
−200
0
200
400
Months from delivery

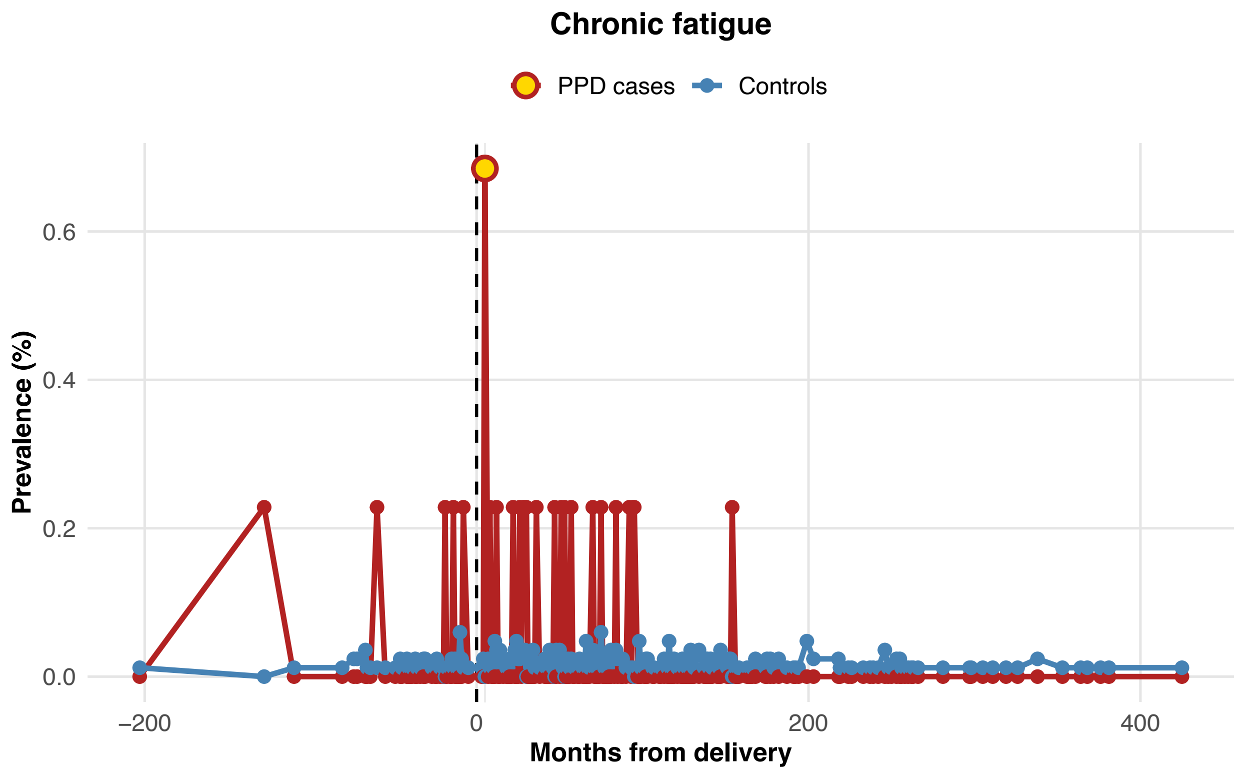
Chronic fatigue
PPD cases
Controls
Prevalence (%)
0.0
0.2
0.4
0.6
−200
0
200
400
Months from delivery

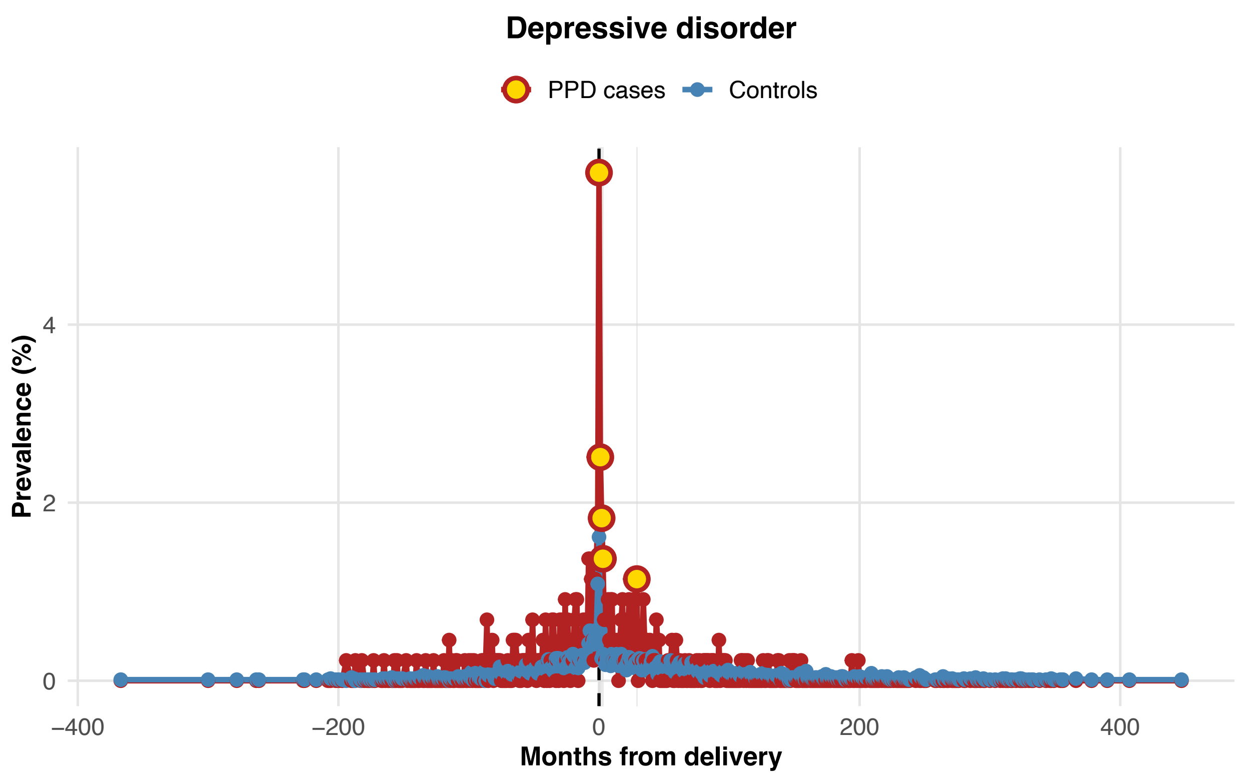
Depressive disorder
PPD cases
Controls
Prevalence (%)
0
2
4
−400
−200
0
200
400
Months from delivery

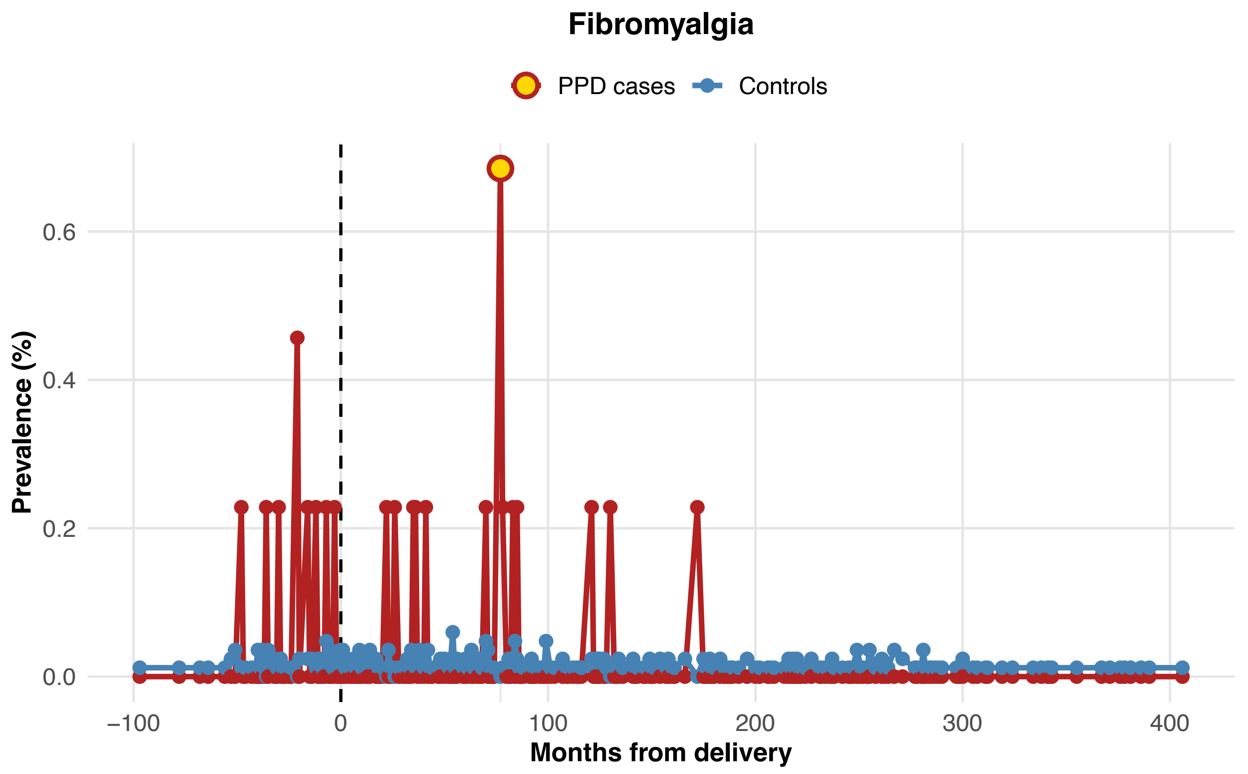
Fibromyalgia
PPD cases
Controls
Prevalence (%)
0.0
0.2
0.4
0.6
−100
0
100
200
300
400
Months from delivery

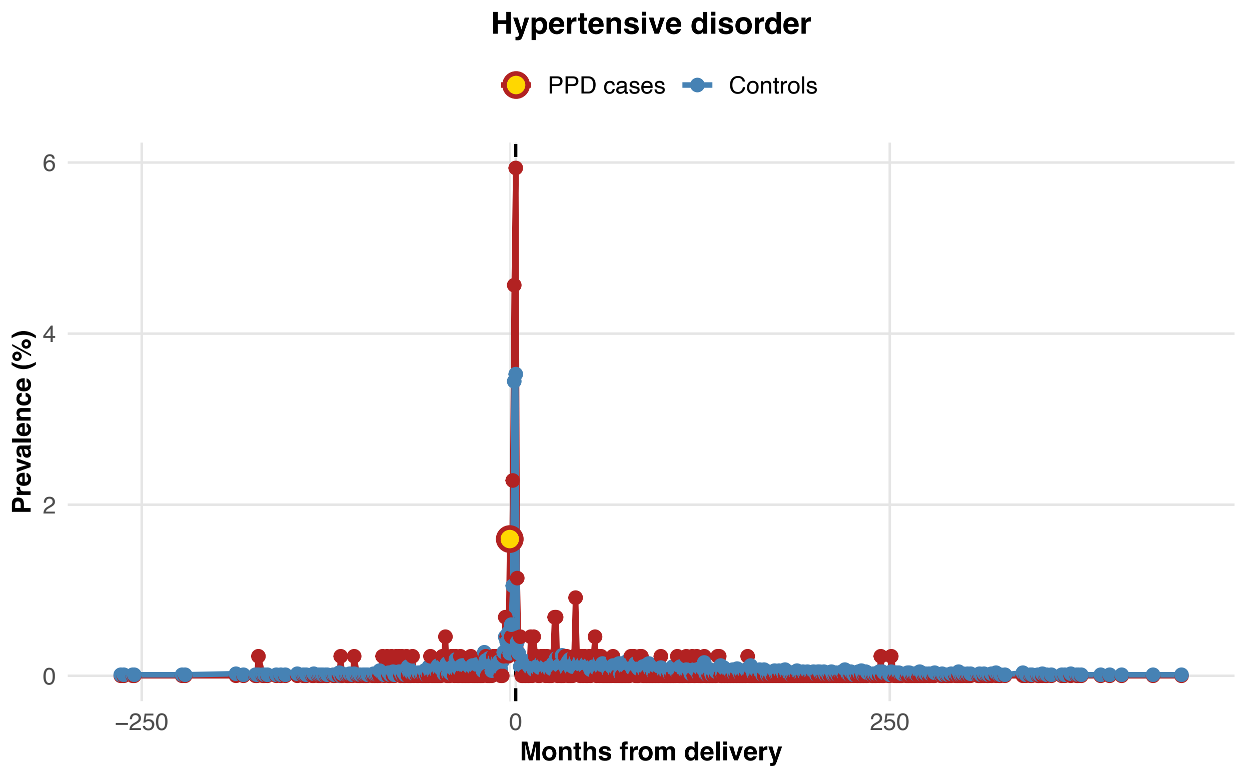

Hypertensive disorder
PPD cases
Controls
Prevalence (%)
0
2
4
6
−250
0
250
Months from delivery


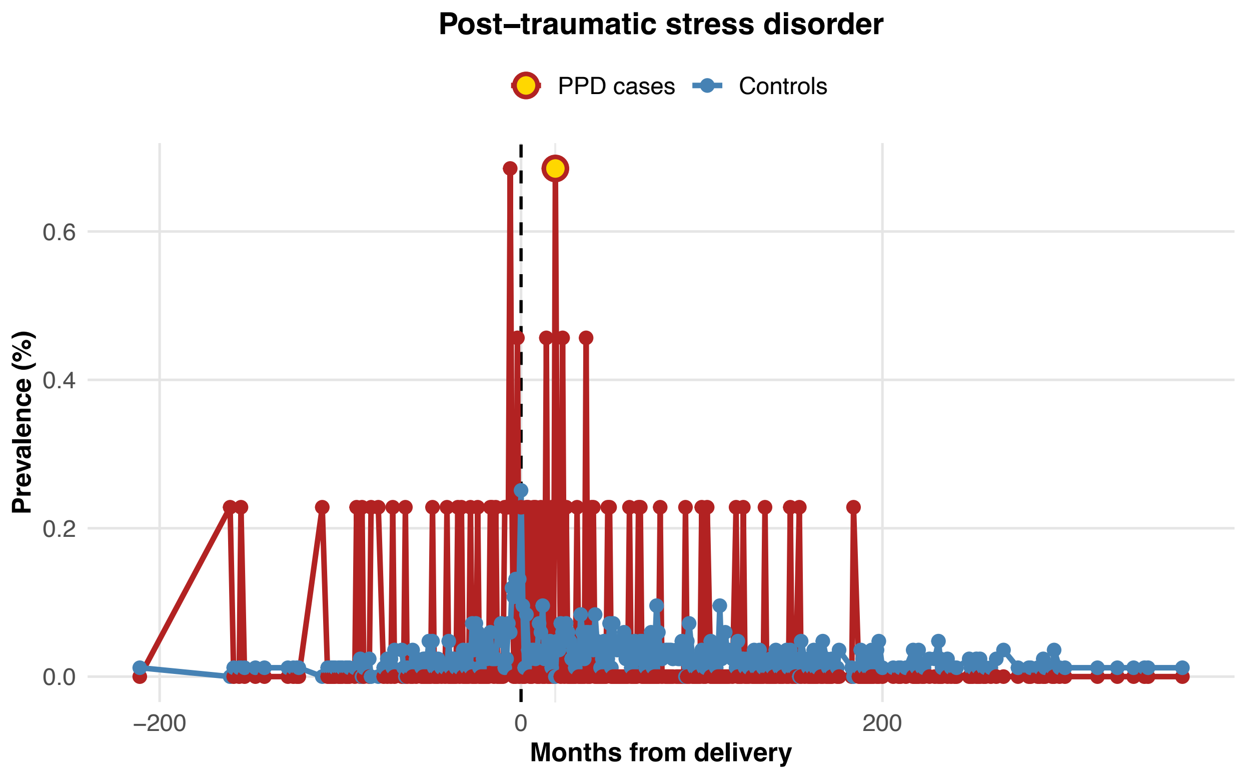

Post−traumatic stress disorder
PPD cases
Controls
Prevalence (%)
0.0
0.2
0.4
0.6
−200
0
200
Months from delivery

Supplemental Figure 2. Correlation between socioeconomic predictors.

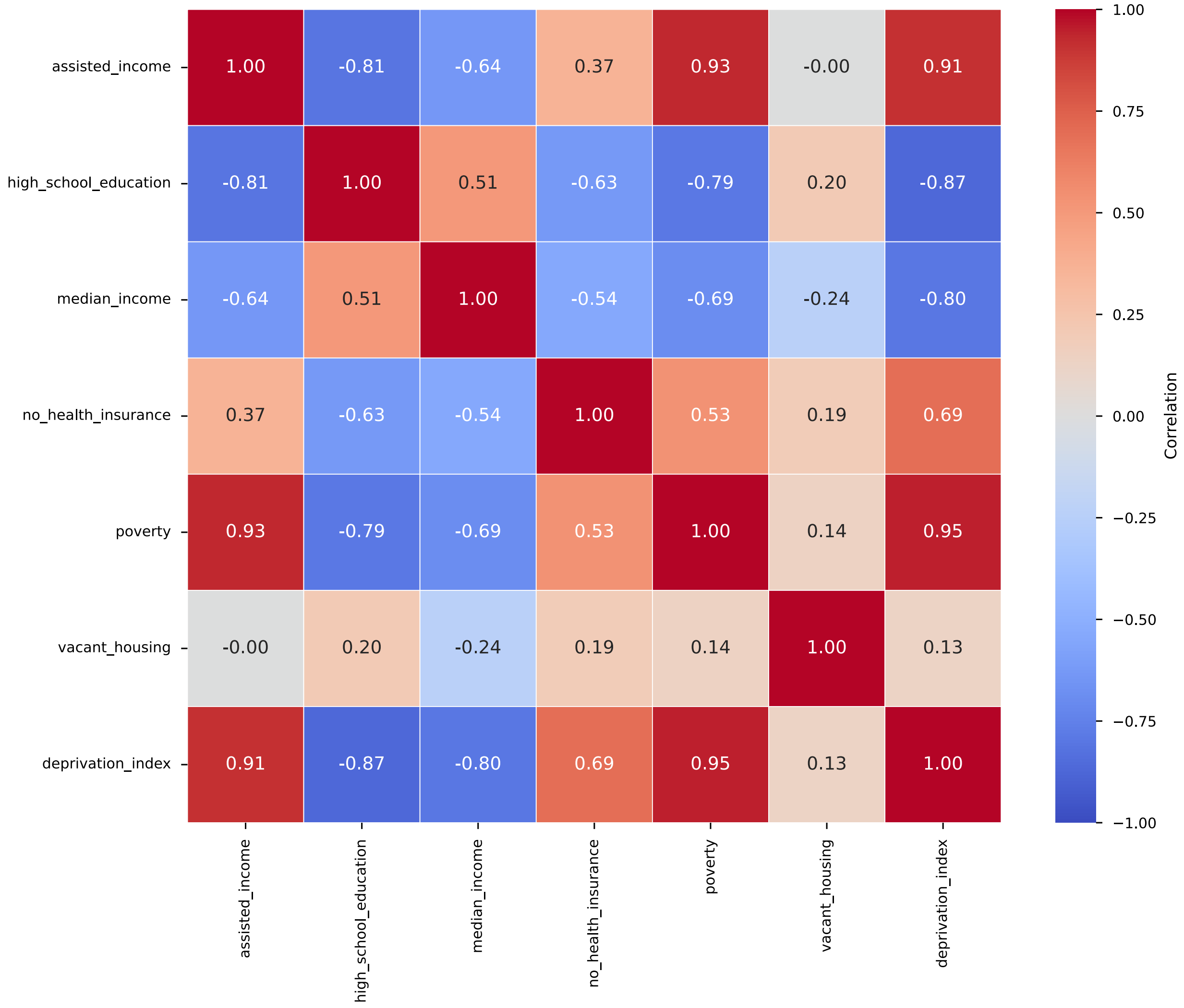

Supplemental Figure 3. Machine learning prediction results for PPD status comparing the performance of three methods, logistic regression, random forest, and XGBoost.

Supplemental Figure 3A. ROC curves.

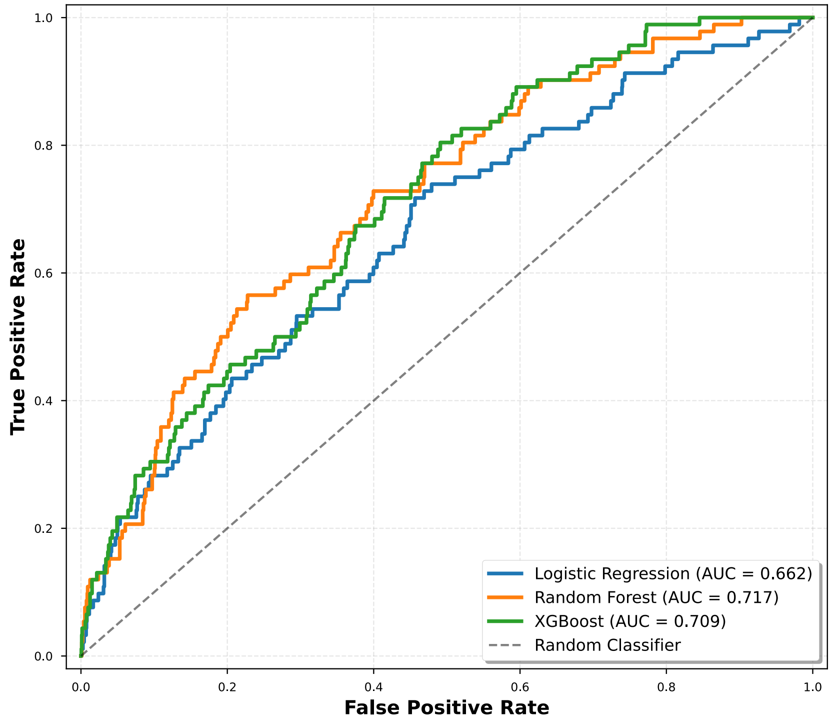


Supplemental Figure 3B. Precision-recall curves.

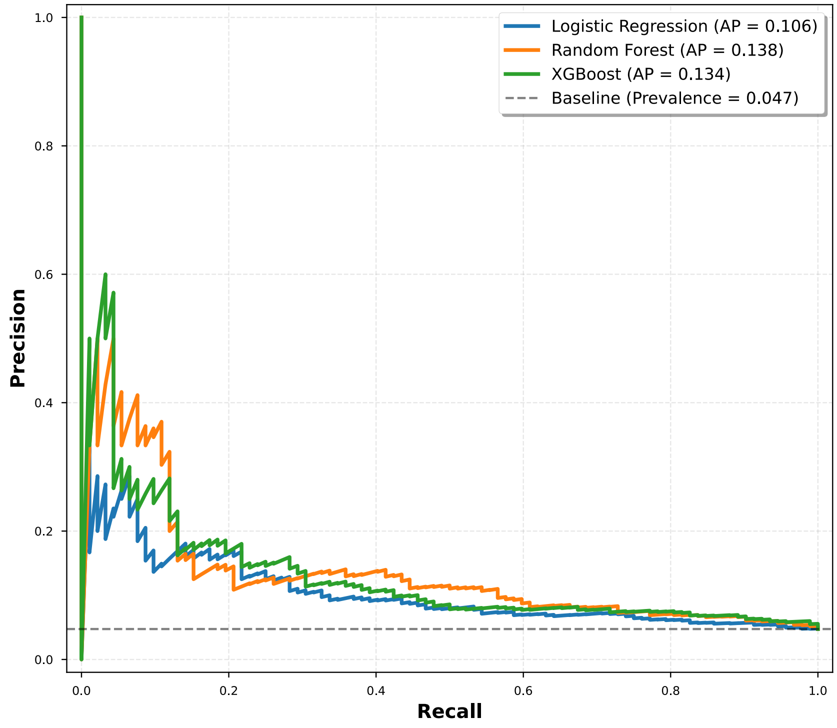

Supplemental Figure 3C. Confusion matrices for the 0.5 threshold.

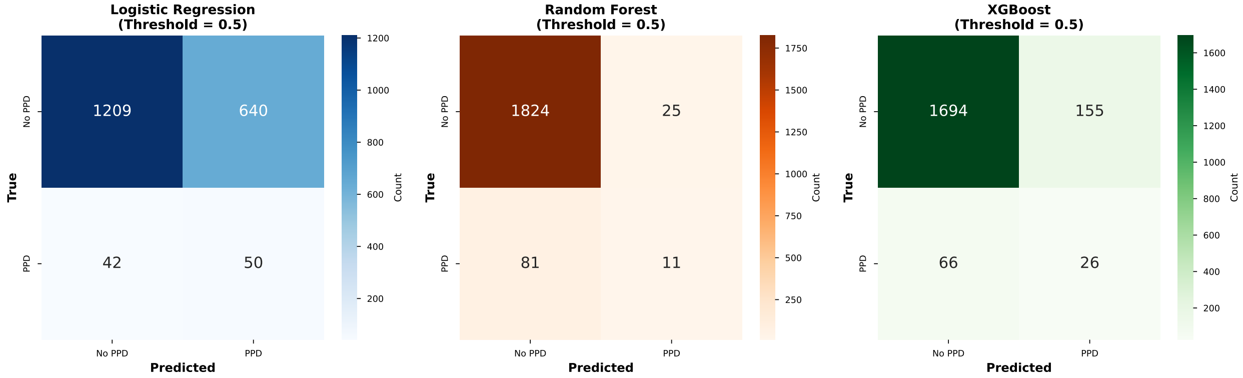


Supplemental Figure 3D. Threshold comparison for the random forest model.

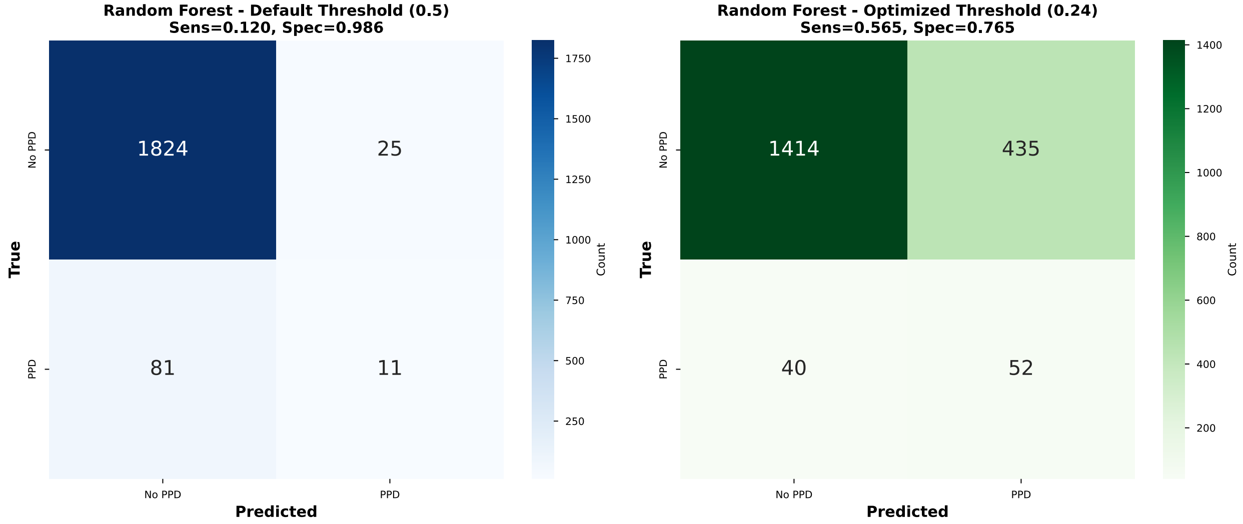

Supplemental Figure 3E. Feature importance.

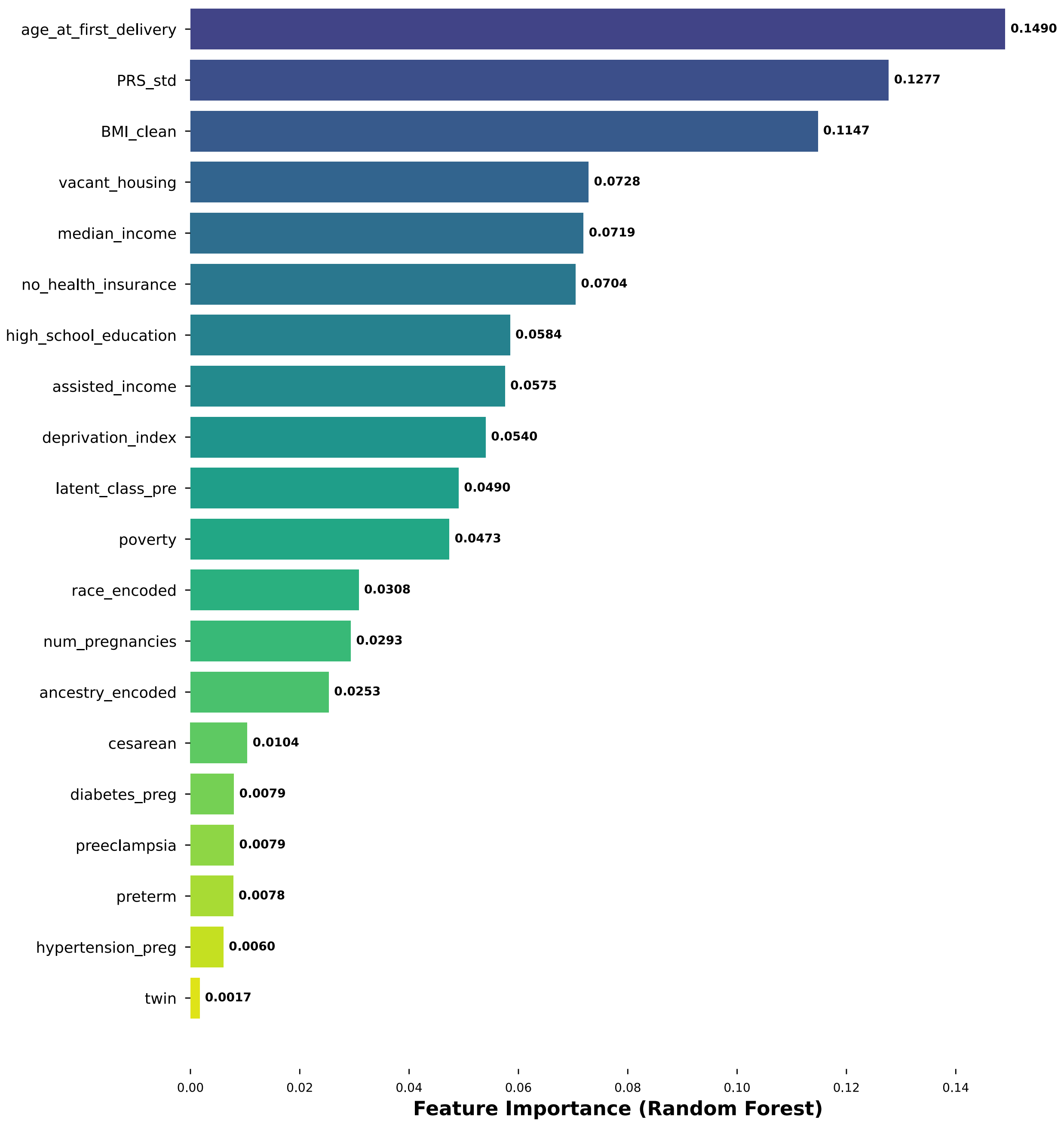

Supplemental Figure 4. Machine learning prediction results for symptomatic post-delivery classes comparing the performance of three methods, logistic regression, random forest, and XGBoost.

Supplemental Figure 4A. ROC curves.

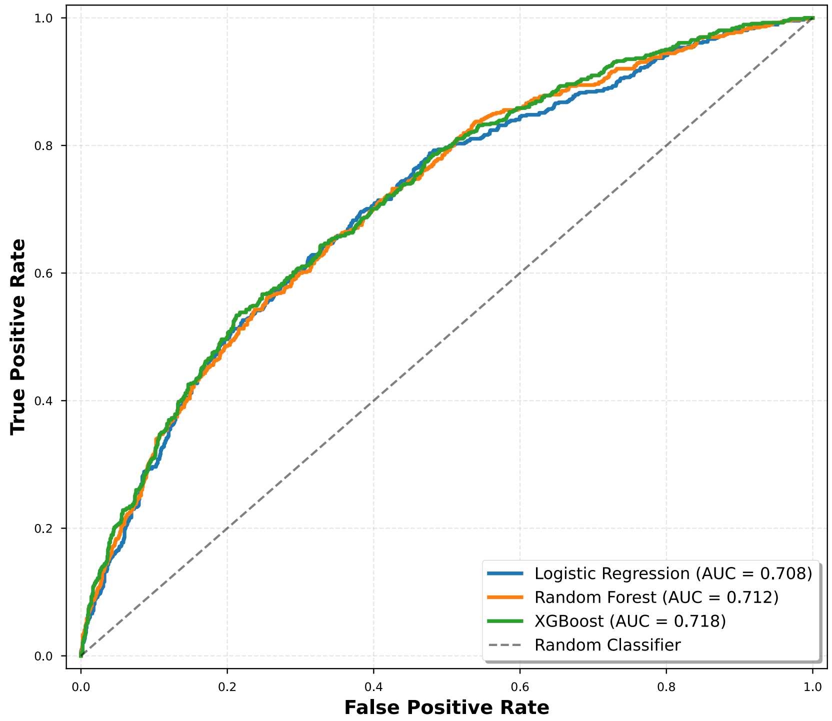


Supplemental Figure 4B. Precision-recall curves.

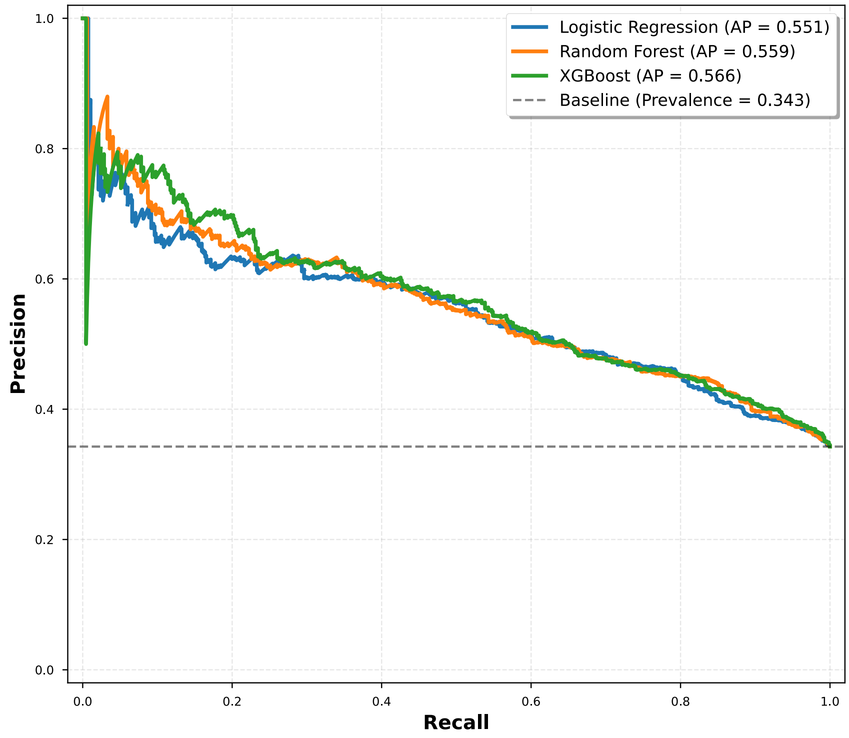

Supplemental Figure 4C. Confusion matrices for the 0.5 threshold.

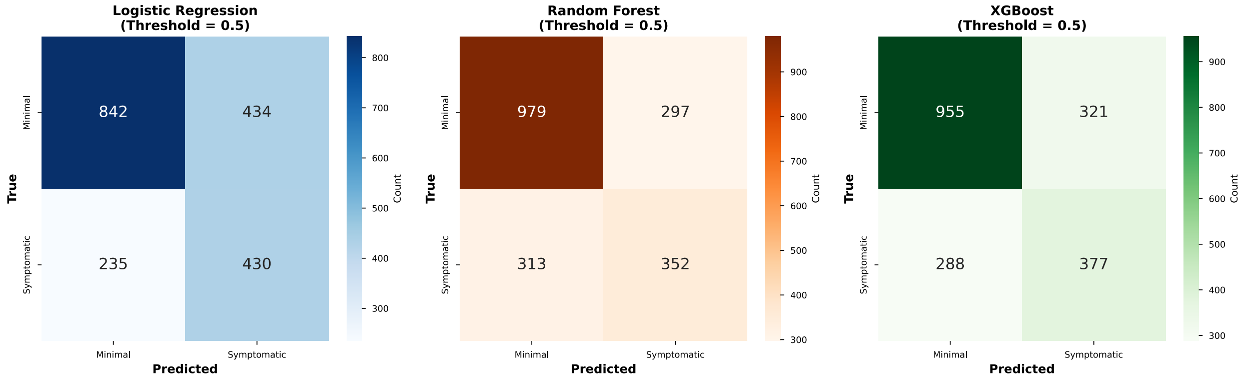


Supplemental Figure 4D. Threshold comparison for the XGBoost model.

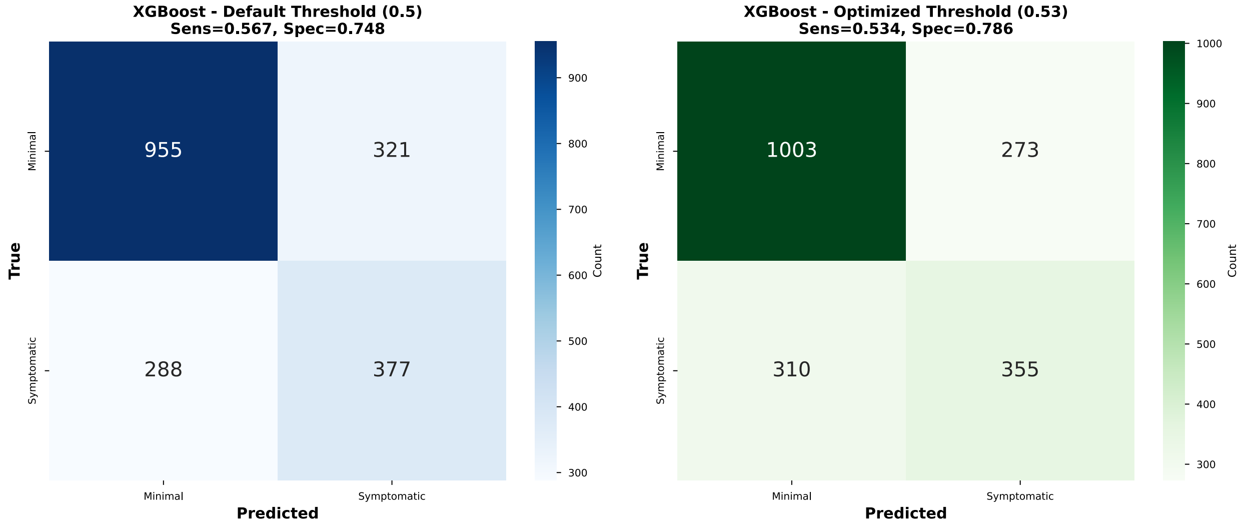

Supplemental Figure 4E. Feature importance.

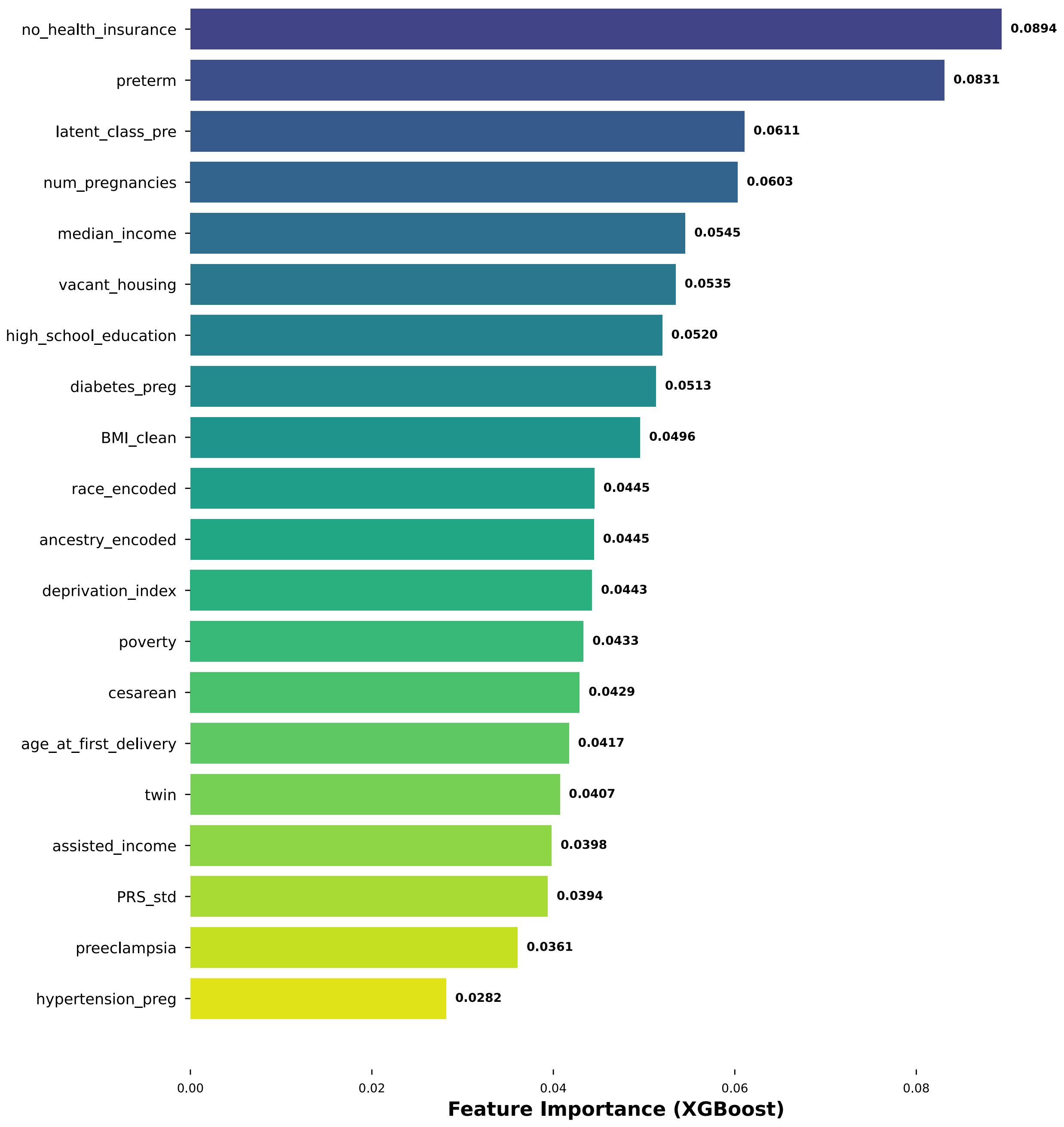

Supplemental Figure 5. Machine learning prediction results for symptom worsening (class transitions from class 1 to symptomatic classes 2 and 3) comparing the performance of three methods, logistic regression, random forest, and XGBoost.

Supplemental Figure 5A. ROC curves.

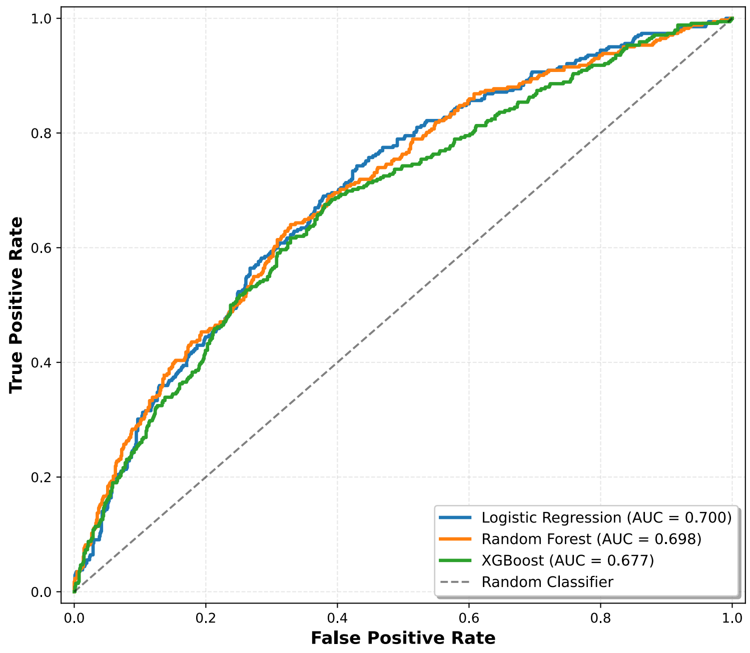


Supplemental Figure 5B. Precision-recall curves.

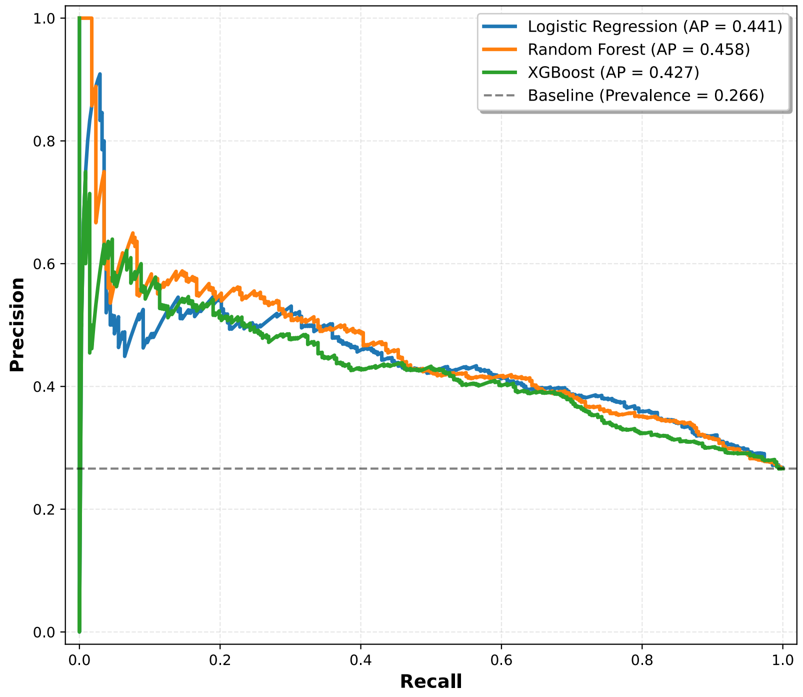

Supplemental Figure 5C. Confusion matrices for the 0.5 threshold.

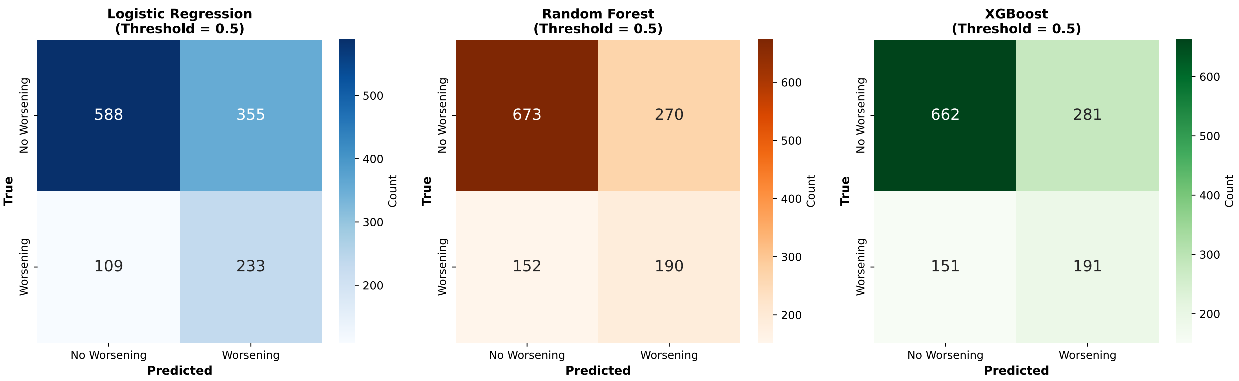


Supplemental Figure 5D. Threshold comparison for the logistic regression model.

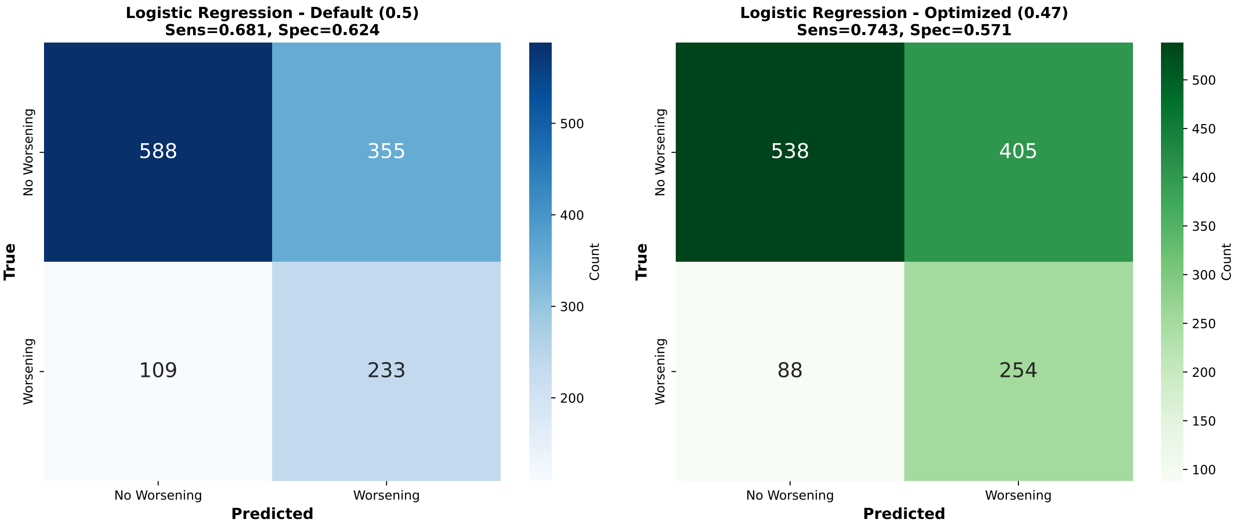

Supplemental Figure 5E. Coefficients.

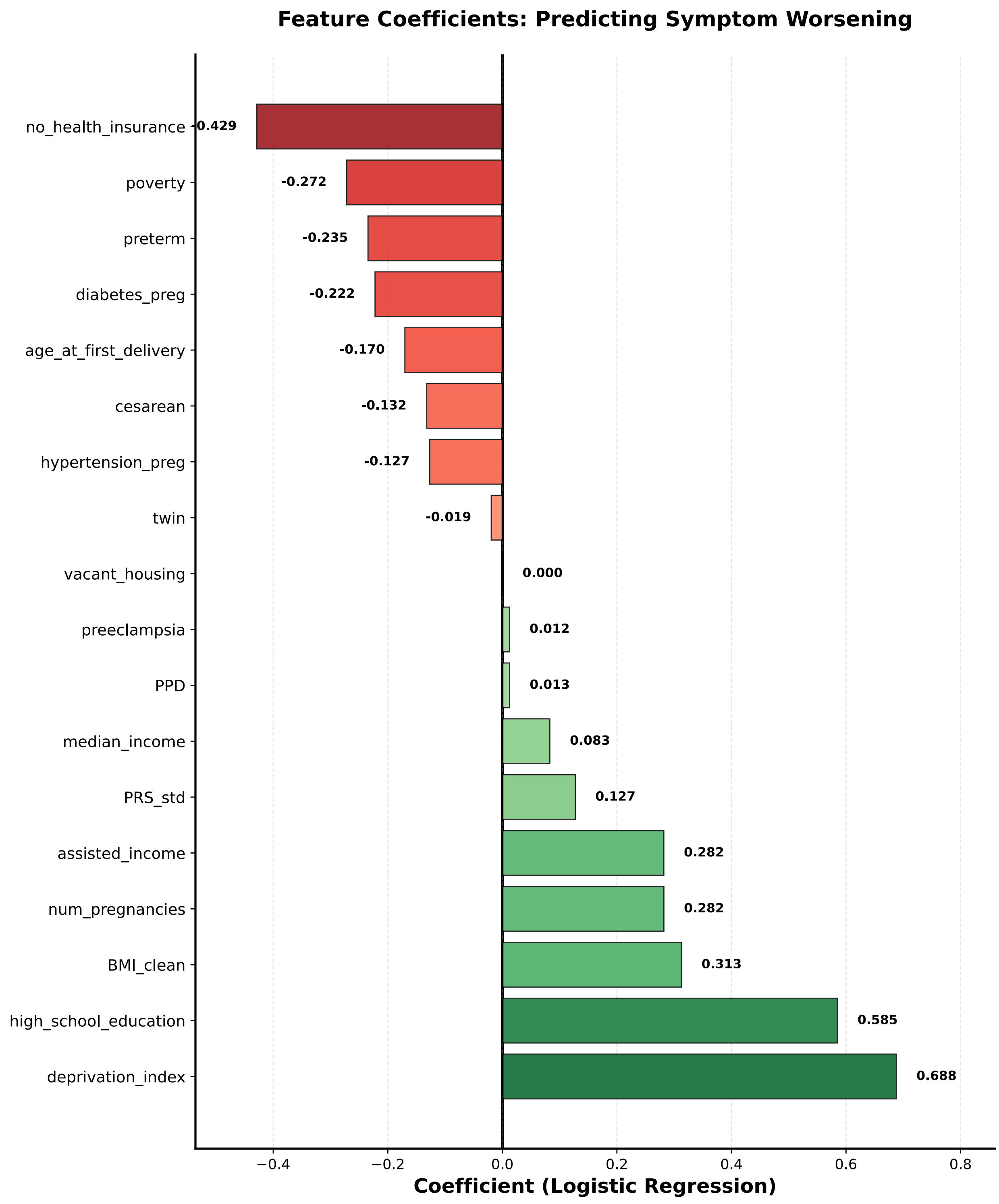
